# Integrating multi-omic QTLs and predictive models reveals regulatory architectures at immune related GWAS loci in CD4+ T cells

**DOI:** 10.64898/2026.01.27.26344979

**Authors:** Marliette R. Matos, Samuel Ghatan, Sean Bankier, Taylor V. Thompson, Kassidy Lundy-Perez, Masako Suzuki, Reanna Doña-Termine, Jacob Stauber, David Reynolds, Kathleen Rosales, Anthony Griffen, Mariko Isshiki, Danny Simpson, Nathanael Andrews, Omar Ahmed, Samantha Gold, Sophia R. Ostrowiak, Srilakshmi Raj, Sofiya Milman, Tuuli Lappalainen, John M. Greally

**Author notes:** These authors jointly supervised this work (Tuuli Lappalainen, John M. Greally).

## Abstract

Functional interpretation is essential for understanding how genetic variants contribute to complex traits. Here, we identified and characterized regulatory variants in CD4+ T cells collected from 362 donors. We integrated molecular QTL mapping from single-cell RNA-seq profiles and chromatin accessibility with predicted variant effects from a deep learning model trained on chromatin accessibility data. We identified molecular features and transcription factor binding mechanisms underlying variant sharing and mediated effects across the modalities and approaches. While predicted variant effects correlated with molQTLs, only a small fraction of empirically detected molQTLs were discovered by predictive models. MolQTLs, primarily those affecting chromatin, indicated potential molecular drivers for 33% of immune-related GWAS loci, with the deep learning approach providing insights into 4.7% of GWAS loci. These results highlight the value of multi-omic data and systematic integration of empirical and predictive approaches to interpret regulatory effects of genetic variants.

## Introduction

Functional interpretation of disease-associated genetic variation remains a major challenge, despite rapid advances in variant discovery and prioritization. Genome-wide association studies (GWAS) have catalogued an ever-growing number of loci associated with complex diseases and traits, yet association signals typically provide limited mechanistic insight beyond genomic location and nearby genes. In addition, most GWAS loci map to noncoding regulatory regions of the genome ^1,2^, are frequently pleiotropic, and individually contribute small effect sizes, leaving many mechanistic associations unresolved.

Quantitative trait locus (QTL) mapping has improved the interpretation of GWAS variants by linking DNA variation to quantifiable molecular phenotypes, providing empirical evidence for the regulatory action of disease-associated variants within molecular systems. Expression QTLs (eQTLs) are the most widely identified type of molecular QTLs and have played a central role in understanding genetic regulation ^3,4^. These analyses have demonstrated that variant effects are often context specific, at times selectively acting in cell types, or transitional states ^5–9^, but also that eQTLs alone cannot capture all the regulatory variation in GWAS ^10^. Therefore, broadening molecular QTL (molQTL) detection from bulk tissue gene expression to purified and single cell systems ^11–13^ and to other quantitative molecular phenotypes such as chromatin accessibility, DNA methylation, etc., has helped better explain the regulatory intricacies of disease-associated variants ^14,15^.

Because molQTL discovery relies on association testing in naturally occurring human variation, its ability to identify variants affecting gene regulation is fundamentally limited by the structure of the data, particularly linkage disequilibrium (LD), allele frequency, and available sample size. Collectively, these factors make it difficult to pinpoint the truly causal regulatory variant(s), a challenge that is most acute for lower-frequency alleles. In parallel, sequence-based deep learning models that infer regulatory activity directly from DNA sequence have emerged as promising tools for variant assessment ^16–19^, enabling variant-level scoring without requiring large cohorts. These approaches make it increasingly feasible to evaluate context-specific regulatory potential of individual variants. However, it remains unclear to which extent such predictions can be interpreted independently of association evidence, and how they should be weighed relative to molQTL fine-mapping for variant prioritization and biological interpretation, particularly at disease-associated loci.

To address these challenges in the context of immune-mediated disease, we generated a multi-omic resource comprising single-cell gene expression profiles and bulk chromatin accessibility measurements in peripheral CD4⁺ T cells from 362 genotyped Ashkenazi Jewish individuals. Using this resource, we mapped cis-eQTLs, dynamic (cell-state dependent) eQTLs, and chromatin accessibility QTLs (caQTLs) within the CD4⁺ T cell compartment. We further performed statistical colocalization of eQTLs and caQTLs and applied causal mediation analysis to infer causality and regulatory directionality, linking genetic effects across molecular layers within the transcriptional regulatory landscape.

In parallel, we trained ChromBPNet models^16^ on our CD4⁺ T cell chromatin accessibility data to determine the contribution of DNA sequence to regulatory activity. We integrated these predictions with our molQTLs to assess the concordance between these fundamentally different approaches, prioritize putative regulatory variants, predict transcription factor motif disruption events, and assess how sequence-based predictions relate to traditional regression-based QTL mapping in the interpretation of disease-associated variation. Finally, we extend our framework to interpret genetic loci implicated across a spectrum of immune diseases and traits, placing fine-mapped GWAS variants into CD4⁺ T cell-specific molecular and regulatory context.

Together, our findings provide cellular and molecular context for numerous GWAS loci and demonstrate that integrating gene expression with chromatin accessibility, and, more broadly, additional molecular phenotypes, can help resolve functional regulatory relationships that improve the interpretation of disease-associated noncoding variation. We also show that function-to-sequence models such as ChromBPNet are most informative when used in a complementary manner alongside molQTL evidence for variant prioritization and molecular interpretation at disease loci.

## Results

### Cohort Characteristics

We generated a comprehensive multi-omic map comprised of genotypes, single-cell gene expression, and bulk-level chromatin accessibility profiles of CD4^+^ T cells purified from the peripheral blood of 408 initially recruited self-identified Ashkenazi Jewish individuals (**Figures S1A-B**). From the initial cohort, genomic DNA from 397 individuals were successfully genotyped, and following quality control, 377 unrelated individual genotypes were retained (kinship cutoff =0.884) along with 7,824,658 imputed variants with minor allele frequency (MAF) greater than 0.05 (See Methods, **Figure S2A**). In parallel, we generated libraries for single-cell RNA sequencing (scRNA-seq) and Assay for Transposase-Accessible Chromatin with sequencing (ATAC-seq) from each individual across 34 experimental batches (**Figures S1D-E**). For each batch, scRNA-seq libraries were multiplexed by pooling CD4^+^ T cells from 12 genetically distinct samples at a time, capturing cells from a total of 398 individuals, while bulk ATAC-seq libraries were prepared independently for each individual, resulting in 397 distinct chromatin accessibility profiles.

For all downstream genetic mapping analyses in this study, the resulting scRNA-seq and bulk ATAC-seq datasets were filtered to include only 362 individuals common across all three sequencing modalities (**Figure S2B**). The final dataset included expression of 26,681 genes from 475,552 cells (median 1,310 cells/individual, 1,492 genes/cell) and 71,348 accessible chromatin peaks (mean depth: 59.9 million paired reads/individual).

### Molecular QTLs explain distinct regulatory contexts across gene expression and chromatin accessibility

To identify genetic variants associated with allelic differences in gene expression and/or chromatin accessibility in CD4^+^ T cells, we mapped cis-eQTLs and cis-caQTLs by testing variants within ±1 megabase (Mb) of a gene’s transcription start site (TSS) or to the summit of an accessible chromatin peak, respectively (**Figure 1A**). At a 5% false discovery rate (FDR), we identified 10,424 conditionally independent cis-eQTLs and 68,294 caQTLs regulating 6,790 unique eGenes and 37,114 unique caPeaks, respectively (**Figure 1B**). Of these, 7,943 unique eQTLs (76.2%) and 42,670 caQTLs (62.7%) were fine-mapped, which were used for most downstream analysis hereon referred to as *fine-mapped molQTLs* (**Figure 1B**).

**Figure 1.**
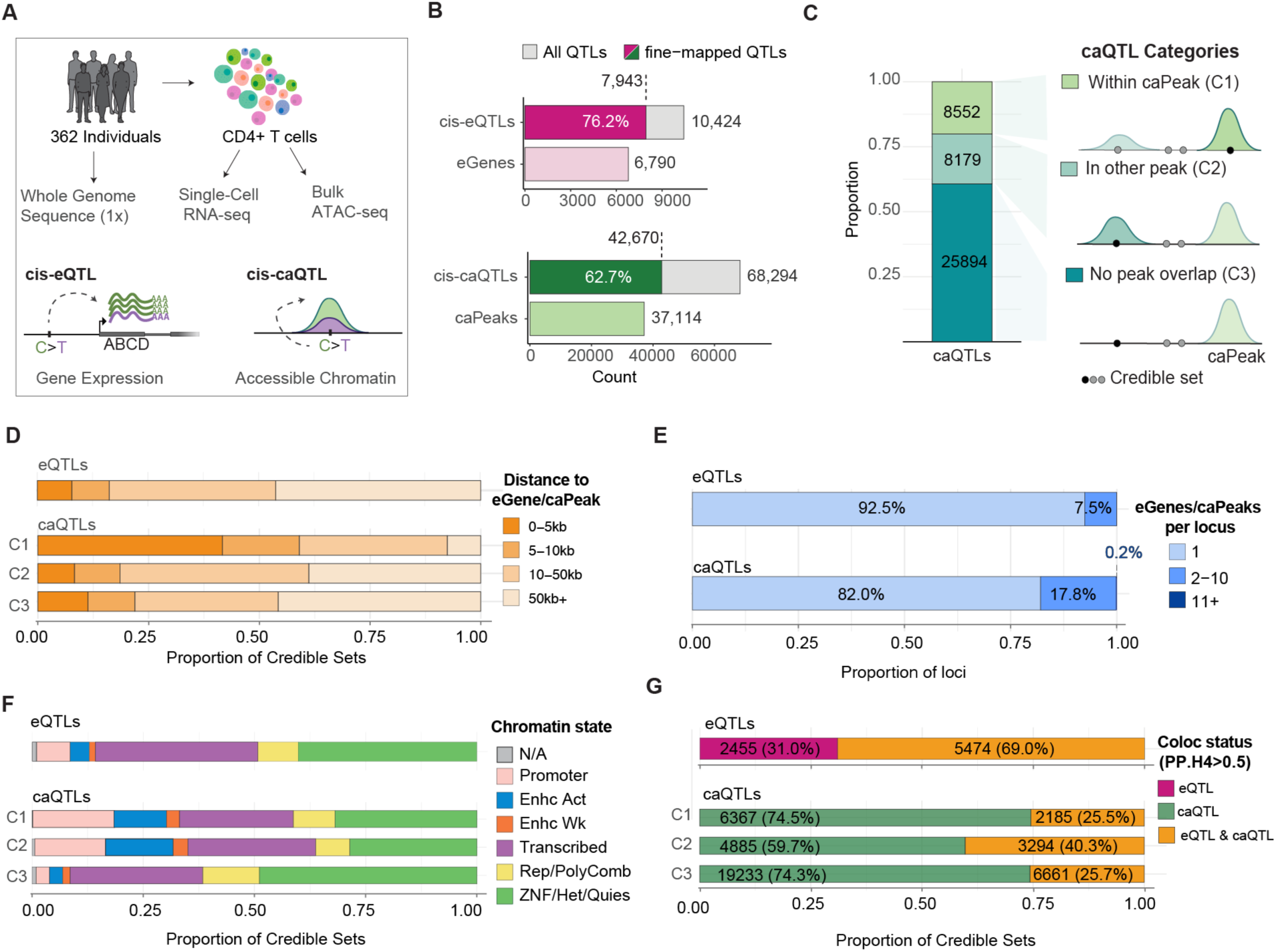
molQTLs differentially predict regulatory architecture across molecular phenotypes. (**A**) Experimental design and overview of molQTL mapping. (**B**) Number of eQTLs/eGenes and caQTLs/caPeaks mapped, shaded by the fraction of loci that were fine-mapped in each QTL category. (**C**) Number of caQTLs per category with a schematic of category definitions. (**D**) Mean distance from each fine-mapped credible set to its associated feature (eGene TSS or caPeak summit), binned by distance. (**E**) Distribution of features (eGenes and caPeaks) associated with eQTL/caQTL loci, binned by the number of features per locus. (**F**) Proportion of chromatin states associated with molQTL CSs, stratified by caQTL categories. (**G**) Proportion of credible sets colocalized between eQTLs and caQTLs, further stratified by caQTL categories.

Variants within open chromatin are more likely to have direct functional effects than variants outside accessible regions^2^. Therefore, we categorized fine-mapped caQTLs into three CS-level categories defined by the highest-priority peak overlap among variants in the credible set (C1 > C2 > C3; Figure 1C; Methods), similar to studies ^6,20^. Overall, 8,552 caQTLs (20.1%) had at least one CS variant overlapping the associated caPeak (C1), 8,179 (19.2%) had at least one CS variant overlapping a different accessible chromatin peak (C2), and 25,894 (60.7%) had no CS variant overlapping any accessible chromatin peak (C3, **Figures 1C, S3A-C**). Due to the strict peak calling criteria (See Methods), not all of these variants are in truly inactive chromatin and some are in areas flanking peak summits (**Figure S3-C).** However, this classification provides a framework for interpreting whether caQTL effects could be mediated by direct disruption of DNA binding sites within peaks or through alternative regulatory mechanisms.

More than 50% of regulated eGenes lie within ±50Kb of the eQTL variant (**Figure 1D**), similarly to C2 and C3 caQTLs, while C1 caQTLs exhibit the shortest variant-caPeak distance due to their definition (**Figure 1C**). However, among distal caQTLs, the mean relative distance of variants to caPeak in C3 is significantly higher than those in distal in-peak C2 caQTLs (Wilcoxon Test, p=1.18×10^-22^), which could point to distal regulatory mechanisms from inaccessible chromatin regions (**Figure S4A**).

To characterize molecular pleiotropy, we examined fine-mapped QTL loci and quantified how many molecular features (eGenes or caPeaks) each locus regulates (See Methods). cis-eQTL regulation was largely specific: 92.5% are associated with the regulation of a single cis-eGene. In contrast, caQTL loci were substantially more pleiotropic across chromatin features (**Figure 1D**), with 18% linked to more than 1 caPeak (Pearson’s X^2^, p=2.84×10^-78^).

We then looked at functional characteristics of the genomic regions harboring CSs. Within cis-eQTLs, CS variants were primarily distributed across intronic (49.2%) and intergenic (26.2%) regions, with a smaller proportion in promoters (17.8%) and UTRs (4.9%) (**Figure S4C**), consistent with previous literature ^21^. Because chromatin accessibility is reflective of a wider net regulatory activity across genomic contexts compared to gene expression, we intersected eQTLs and caQTL CSs with ENCODE chromatin-state annotations in CD4^+^ T cells, summarized into six major supergroups (**Figure 1F**). Among molQTLs, we did not observe a difference in the proportion of CSs at promoter regions (Pearson’s X^2^, p=0.098). However, eQTLs were strongly represented at actively transcribed regions (Pearson’s X^2^, p=5.1×10^-31^) compared to caQTLs, which were most predominant at enhancers (Pearson’s X^2^, p=1×10^-6^) and repressed chromatin states (**Figure S4D**). Compared to other caQTLs, C1 showed higher enrichment for promoter-associated caQTLs (Fisher’s exact test, OR=4.15, p=9.11×10^-189^), whereas C2 caQTLs followed similar patterns yet were most enriched at active enhancers (Fisher’s exact test, OR=3.79, p=3.71×10^-126^) (**Figures S4E**). However, C3 were strongly enriched at repressed chromatin states compared to other categories (OR=2.19, p=5.21×10^-206^) with up to 61.6 % of CSs at heterochromatin, quiescent, and polycomb-repressed regions (**Figure 1F**).

With these findings in mind, we next examined how eQTLs and caQTLs overlap. To identify molecular QTLs that jointly regulate gene expression and chromatin accessibility, we performed statistical colocalization of eQTLs and caQTLs (PP.H4 > 0.5). We observed that while up to 69% of eQTLs also show evidence of chromatin association, only 30.5% of caQTLs colocalize with eQTLs (**Figure 1G**), highlighting a broad landscape of genetically driven chromatin variation that does not translate into detectable expression changes. Colocalization rates also differed across caQTL categories (Pearson’s X^2^, p =8.42×10^-151^, **Figures S4F**), with the highest proportion in C2 (40.3%), compared to a mean of 25.6% in C1 and C3 caQTLs. Together, these results suggest that molQTLs capture distinct signatures of genetic regulation within the transcriptional regulatory landscape of CD4^+^ T lymphocytes.

### T cell state dynamics modulate genetic effects across immune cellular programs

Given the dynamic nature of the CD4^+^ T cell compartment, encompassing naïve, memory, regulatory, and effector subtypes ^22^, we sought to identify eQTLs modulated by immune cellular states. To identify such events, we used CellRegMap^23^ to test genotype-by-context (GxC) interactions between our previously mapped cis-eQTLs and continuous cellular states, defined by gene expression patterns, in the CD4^+^ T-cell compartment.

To reduce sparsity in the scRNA-seq data and improve detection power, we generated donor-level pseudocells ^23,24^ by clustering transcriptionally similar cells across Leiden resolutions (3.4–5.0; **Figures S5E–F**). We then fit Multi-Omics Factor Analysis (MOFA) models ^25,26^ to the pseudocell expression matrices to identify latent factors capturing major axes of CD4⁺ T cell transcriptional variation. Consistent with prior studies ^22^, baseline expression heterogeneity in unstimulated CD4⁺ T cells was modest (**Figure S5G**). Inspection of factor gene loadings together with GO Biological Process enrichment (hypergeometric test) highlighted continuous state programs spanning naive-to-memory differentiation, stress-response activation, and cytotoxicity (**Figures S5I–J**).

We tested cellular state interactions for 6,939 conditionally independent cis-eQTLs (restricted to one lead signal per gene, see Methods) and identified 210 cis-eQTLs whose effect sizes vary significantly in CD4⁺ T pseudocells along transcriptional trajectories. Figures 2A-B show examples of dynamic cis-eQTLs for Interleukin 32 (IL32) and granzyme M (GZMM), where genotype-dependent expression is progressively modulated as CD4+ T cells transition between immune states.

**Figure 2.**
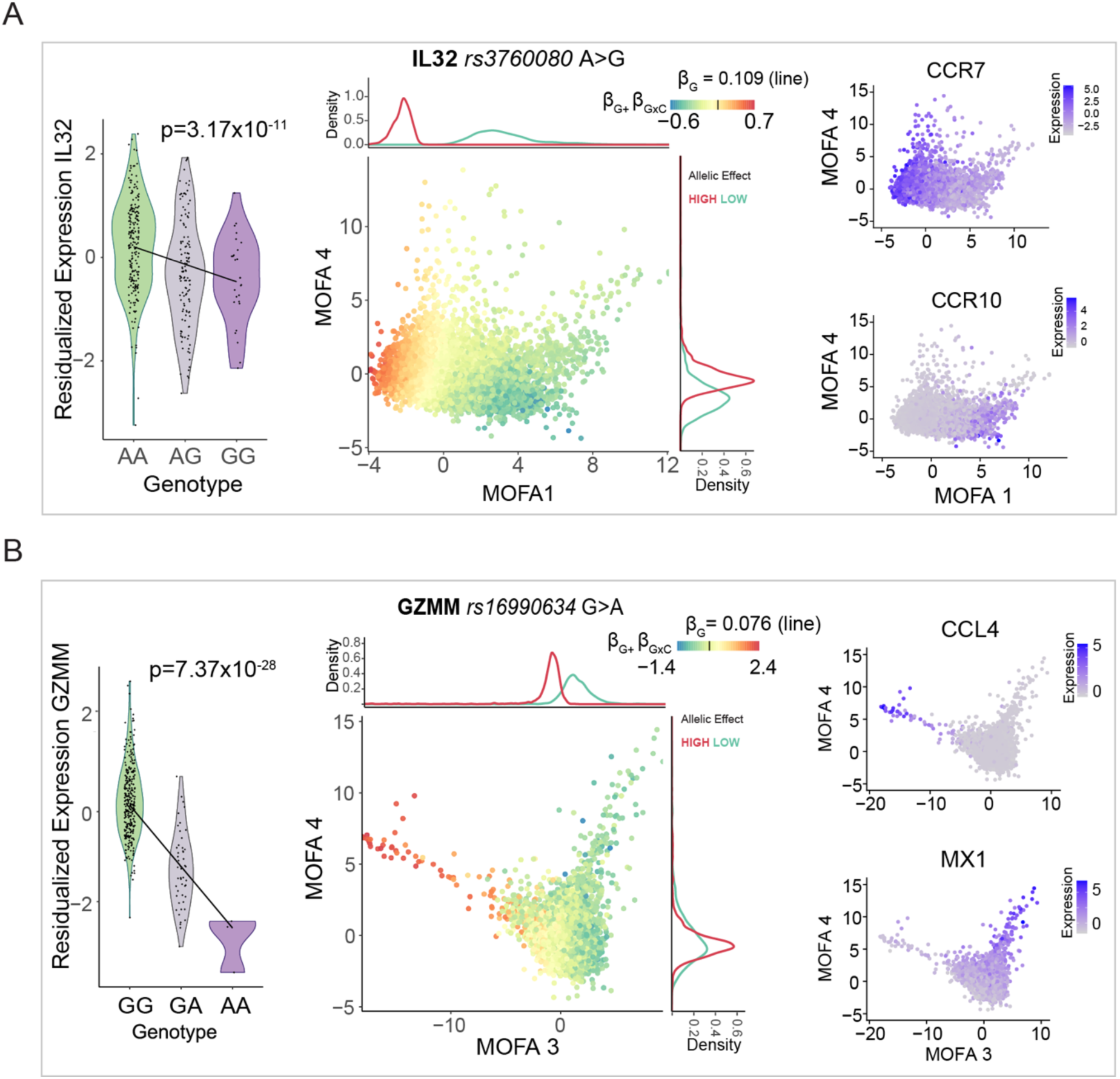
Genetic regulation is dynamically modulated by immune cellular states. Representative examples of cellular state-dynamic cis-eQTLs for (**A**) *IL32* and **(B**) *GZMM* respectively. (*Left*) violin plots of residualized pseudo bulk expression of each eGene stratified by eQTL genotype, BH corrected p-values. (*Middle*) MOFA embedding of pseudocells colored by the genotype effect as a function of cellular state (allelic effect, β_G_ + β_G×C_). The color legend is centered on the overall genetic effect (β_G_, vertical line), and density plots on the sides show where pseudocells with strongly increased or decreased allelic effects (top/bottom 10%) fall along the trajectory. (*Right*) MOFA projections of pseudocells colored by expression of marker genes defining the cellular state.

The eQTL for IL32 (Tag SNP: rs3760080 A>G) is associated with decreased expression of IL32 (p=3.17×10^-11^) for individuals carrying the alternative allele (ALT). We observed that the genetic effect associated with rs3760080 on IL32 is most prevalent among naive CD4^+^ T cells. This is most evident across pseudocells projected on Factor 1, which captures a naïve-to-effector CD4^+^ T cell differentiation axis, marked by decreasing expression of the lymph node-homing receptor CCR7 ^27,28^ and strong positive loadings for the tissue-homing receptor CCR10 ^29^ and activation markers including HLA-DR genes (**Figures 2A, S5I**). The progressive change in expression of these markers illustrates the gradual polarization of naïve T cells to differentiated states. IL-32 has been implicated in inflammatory responses and autoimmune disease ^30,31^. Notably, increased IL32 levels in rheumatoid arthritis have been associated with rs4786370 ^32,33^, which is in linkage disequilibrium (R^2^=0.2719, p<0.001, Pearson’s X^2^) with rs3760080 (**Figure S6A**, See Methods). Collectively, our data highlight baseline (naïve) CD4⁺ T cell regulation as a candidate context for interpreting IL32-associated regulatory variants reported in rheumatoid arthritis studies.

Similarly, GZMM encodes granzyme M, a cytotoxic effector protease released from lytic granules during target-cell killing ^34^. Although cytotoxicity has been historically framed as a CD8⁺ T cell feature, CD4⁺ T cells can also become cytotoxic, especially with chronic activation and with age ^35–38^. In our data, the eQTL effect on GZMM is strongest in a cytotoxic/inflammatory cellular program, defined by the co-expression of CCL4, GZMB, CX3CR1, PRF1 ^35^ (**Figures 2B, S5I-J)**. Since the cis-eQTL (tag SNP rs16990634 G>A) is associated with decreased GZMM expression in alternative-allele carriers (p=7.37×10^-26^) (**Figures 2B**), this suggests that those individuals may have reduced GZMM upregulation specifically in cytotoxic-like CD4⁺ states, which could interfere with their immune cytotoxic potential.

### Causal mediation analysis reveals distinct regulatory architectures associated with molQTLs

While colocalization between eQTLs and caQTLs suggests a shared causal variant, it does not establish a regulatory relationship or its direction between the molecular traits. We therefore performed mediation analysis to assess whether allelic effects on chromatin accessibility mediate allelic effects on gene expression, and vice versa (**Figure 3A**); hereafter referred to as the chromatin-to-gene and gene-to-chromatin models.

**Figure 3.**
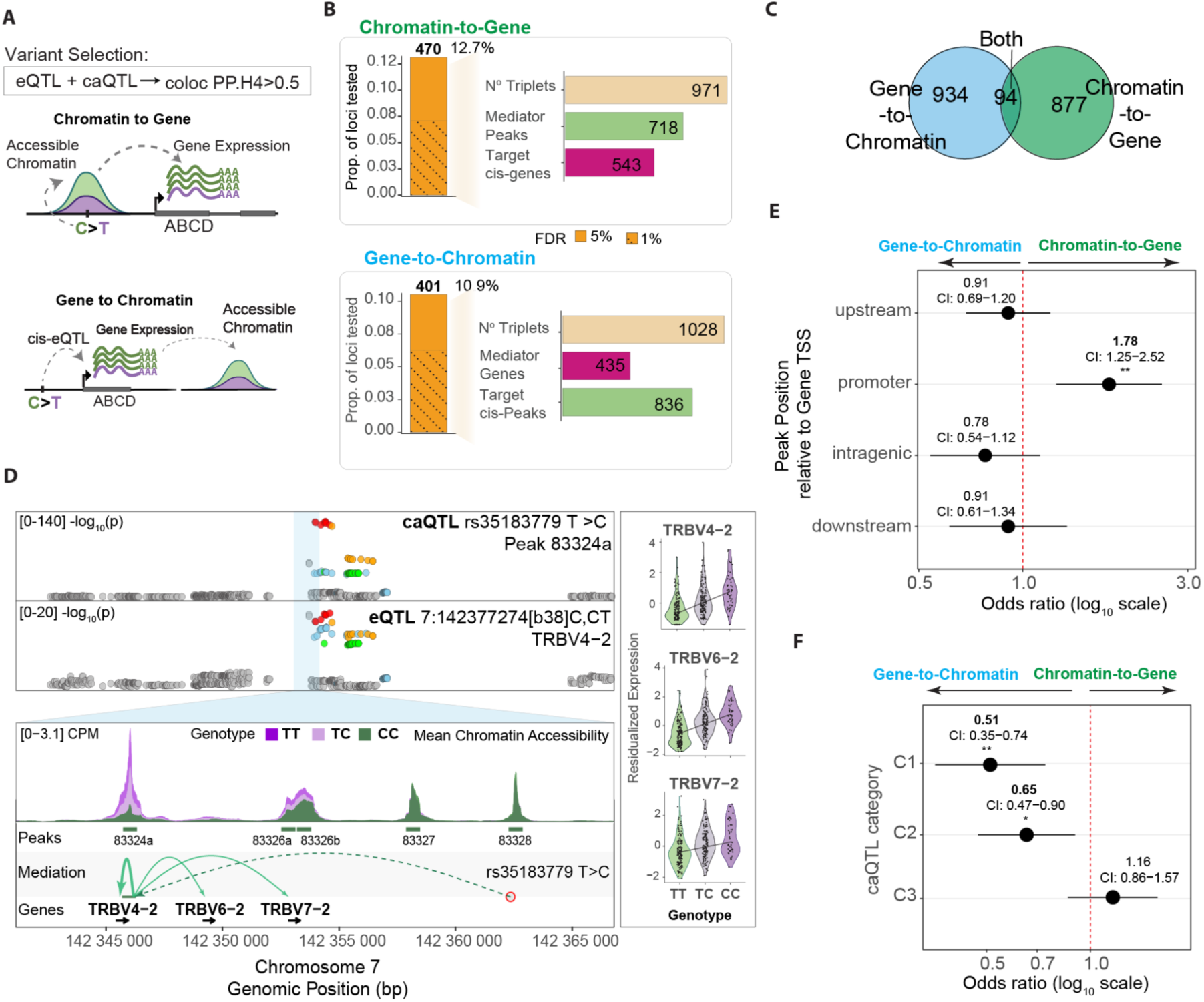
Causal Mediation Analysis of eQTL-caQTL colocalized loci. (**A**) Graphical representation of the causal mediation models tested (*top*) Chromatin-to-Gene and (*bottom*) Gene-to-Chromatin) **(B)** Summary of causal mediation testing for each model. (*Left*) Vertical bar plot quantifying the proportion (y-axis) of loci with significant evidence of mediation via each model with respect to the number of loci tested; (*Right*) Horizontal bar plots quantifying the number of locus-collapsed unique triplets, -mediators and -targets for each model (**C**) Overlap of significant triplets between models. (**D**) Example of chromatin-to-gene mediation event for colocalized caQTL/eQTL loci regulating TRBV4-2 (*top*) locus plot showing colocalization signals, (bottom) mediation signals showing variant to peak regulation (dashed line), and chromatin to cis-genes mediation (solid lines), (*right*) violin plots depicting the residualized expression of TRBV4-2, TRBV6-2, TRBV7-2 per genotype (**E**) Enrichment of caPeak positional classes relative to the eGene TSS across mediation models. For each class (upstream, promoter, intragenic, downstream), we used logistic regression to compare Chromatin-to-Gene versus Gene-to-Chromatin loci (class vs all others), reporting odds ratios (OR) with 95% confidence intervals; p-values were BH-corrected across classes and standard errors were locus-clustered. (**F**) Enrichment of caQTL categories across mediation models, using the same logisticregression framework; ORs (95% CI) are shown with BH-corrected p-values across categories and locus-clustered standard errors..

We used Findr ^39^ to test 18,309 statistically colocalized variant-gene-chromatin peak triplets (PP.H4 > 0.5) for evidence of causal mediation under the two directional models at FDR < 0.05. We identified 470 colocalized genetic loci (12.7 % of collapsed, unique loci tested) with significant support for chromatin-to-gene mediation, spanning 971 triplets (**Figure 3B**). 401 loci (10.9 % of loci tested, 1028 triplets) showed significant support for gene-to-chromatin mediation, consistent with gene expression mediating allelic effects on the accessibility of nearby cis-peaks. Although the number of significant loci was similar across models, gene-to-chromatin mediation mapped to more chromatin targets than chromatin-to-gene mapped to gene targets, as expected given the larger number of peaks tested (71,348) relative to genes (26,681). A small subset of triplets exhibited evidence for mediation under both models (**Figure 3C**), which may reflect more complex regulatory architectures such as feedback loops.

As an illustrative example of chromatin-to-gene regulation, we highlight a colocalized molQTL locus (PP.H4 = 0.945) in which both chromatin accessibility at a nearby cis-peak and the expression of TRBV4-2 share the same underlying genetic signal (**Figure 3D**). After performing causal mediation analysis within this colocalized triplet, we observed strong support for the chromatin-to-gene model, reflected by a high posterior probability (Findr p2p3 = 0.9994) for chromatin-mediated regulation. Consistent with this inference, donor genotypes show allele-dependent accessibility at the cis-peak that tracks with the corresponding allelic differences in TRBV4-2 expression as well as that of neighboring genes TRBV6-2 and TRBV7-2.

To further compare the genomic contexts of the two mediation models, we tested whether chromatin-to-gene and gene-to-chromatin loci differ in their positional and functional annotations (See Methods). We classified each chromatin peak by its position relative to the associated gene (upstream, promoter-proximal, intragenic, downstream) and tested whether each class occurs more frequently in one mediation model than in the other (**Figure 3E**). Promoter-proximal peaks were more frequent in chromatin-to-gene loci (binomial GLM, OR = 1.78, p=0.00546), whereas the other positional classes did not show significant differences between models.

Using the same between-model framework, we next asked whether the types of caQTL variant-caPeak relationships (**Figure 1C**) differ between the mediation models. We found that gene-to-chromatin loci were more frequently associated with caQTL variants that fall within its associated caPeak (binomial GLM, C1, OR = 0.51, p_adj_=0.00110) and those that fall within other peaks (C2, OR = 0.65, p=0.0149), compared to chromatin-to-gene loci (ORs with 5% FDR-corrected P values shown; **Figure 3F**).

Because we tested both models on the same colocalized variant-peak-gene triplets (each predicting both accessibility and expression), differences in support of mediation reflect regulatory architecture rather than different loci. Promoter-enriched chromatin-to-gene loci are consistent with variants acting through discrete elements where accessibility changes mediate transcription. In contrast, gene-to-chromatin loci, where a smaller set of gene mediators maps to many cis-peaks and variants more often fall within peak contexts, suggest accessibility is a coupled downstream readout of transcriptional activity.

### Integrating function-to sequence models with caQTLs to prioritize putative causal variants modulating chromatin accessibility in CD4^+^ T cells

Since most molQTL variants lie in noncoding regulatory regions ^1,2^ (**Figures 1F, S4C**), characterizing how DNA sequence features might shape chromatin accessibility is essential for predicting the functional impact of individual variants. To investigate this, we trained CD4^+^ T cell-specific, bias-factorized ChromBPNet models^16^ using pooled chromatin accessibility profiles (125,944 peaks, 120k mean reads/individual) from 8 individuals in our cohort (**Figures S7A-B**). We then used these trained models (median Pearson r=0.752, **Figures S7C**) to evaluate the allelic effects of all genotyped SNPs within accessible chromatin regions, regardless of their molQTL status, providing a sequence-based prediction of the allelic impact of all candidate variants on CD4^+^ T-cell chromatin accessibility.

We tested the effects of DNA sequence on chromatin accessibility for 159,908 SNPs located within open chromatin regions **(Table S1)**. Following the authors’ recommendations, we limited our analysis to variants within peaks and used the Integrative Prioritization Score (IPS) as the primary measure for variant prioritization^16^, which we refer to as the ChromBPNet score, we identified 11,440 variants that significantly contribute to chromatin accessibility at an empirical p-value_IPS_<0.05, hereafter referred to as ChromBPNet variants. Significant variants showed near-balanced allelic ALT/REF effects on local chromatin accessibility (48.5% positive vs 51.5% negative; binomial test vs 50:50, p = 0.0013, **Figure 4A**), although variants are not independent due to LD. However, ChromBPNet-significant variants were progressively enriched near gene TSSs relative to non-significant variants (Fisher’s exact test, p≤5kb = 3.9×10^-26^, **Figures 4B, S7D**).

**Figure 4.**
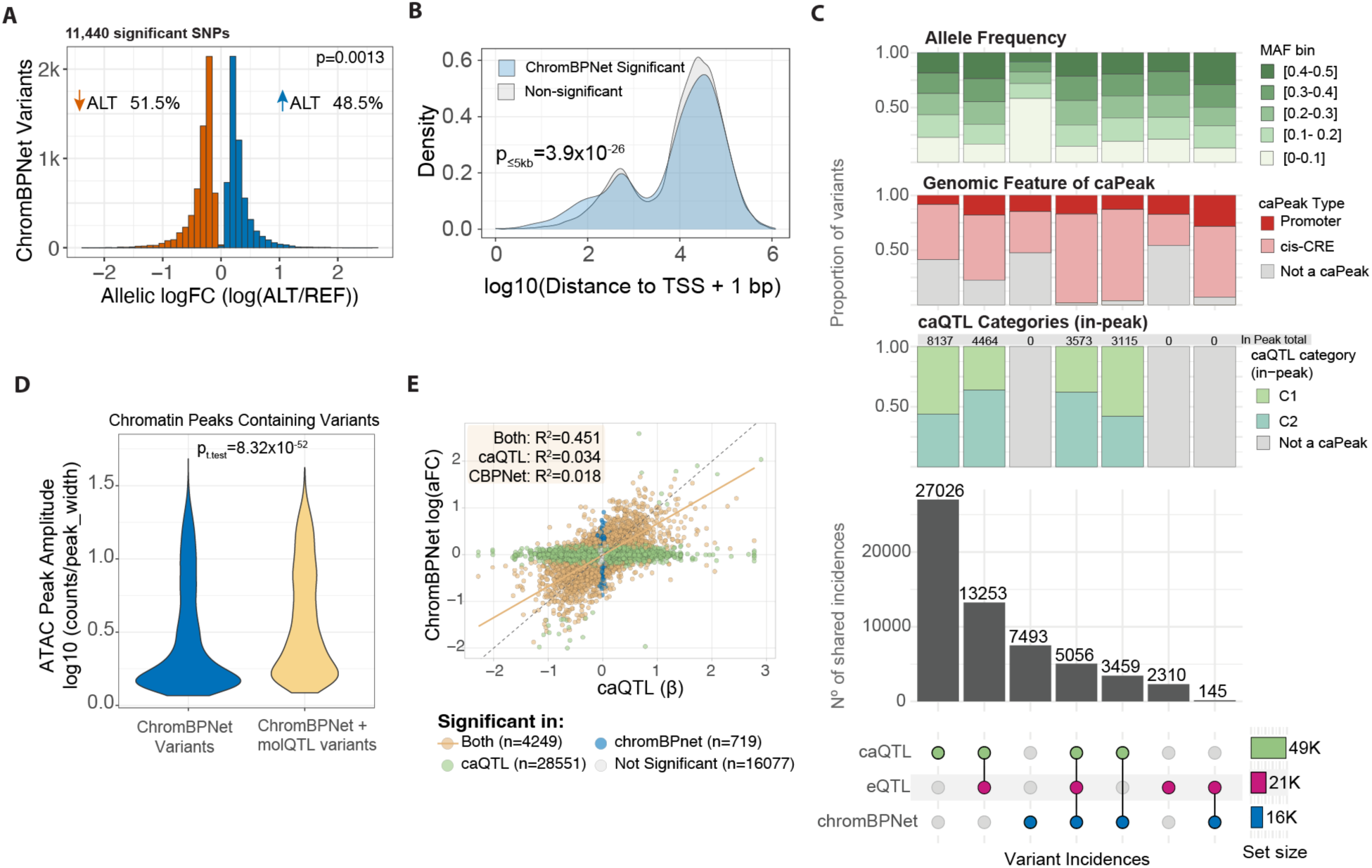
ChromBPNet prioritizes candidate causal variants within molQTL credible sets. **(A)** Distribution of the ChromBPNet Allelic logFC predicted for significant variants (p_IPS_<0.05), colored by ALT allele effect on chromatin accessibility (red) negative and (blue) positive. **(B)** Density plot of distance to the nearest transcription start site (TSS), plotted as log10(distance + 1 bp), stratified by IPS significance based on empirical P values (blue, significant; gray, not significant). **(C)** UpSet plot showing intersections between ChromBPNet-prioritized variants and molQTL credible sets (CSs) containing at least one ChromBPNet variant. Stacked barplots summarize, for each intersection set, the proportions (*top*) of variants within minor allele frequency (MAF) bins, (*middle*) overlap with caPeak genomic annotations (*bottom*) variants within in-peak caQTL categories (**E**) Violin plot of the distribution of ATAC-seq read coverage at peak-containing ChromBPNet variants, stratified by variants specific to ChromBPNet and those shared between ChromBPNet and molQTLs. (**F**) Scatter plot comparing ChromBPNet allelic logFC and caQTL regression beta for variants tested in both methods.

We then assessed the concordance between ChromBPNet-based predictions and caQTL mapping in identifying genetic variants that affect chromatin accessibility and evaluated the predictive value of these variants for gene expression. We quantified overlap by treating each molQTL CS as a unit and defining overlap as the presence of at least one ChromBPNet-significant variant within the CS. Upon intercepting ChromBPNet significant variants with CSs of caQTLs and eQTLs (**Figure 4C**), we observe the highest overlap of ChromBPNet variants within colocalized caQTL/eQTLs (5056 shared incidences), followed by 3459 shared variant incidences between ChromBPNet and caQTLs, with the smallest overlap between ChromBPNet variants and eQTLs, sharing only 145 variant incidences. Given that ChromBPNet is trained on chromatin accessibility profiles, higher representation within caQTLs and caQTL-predicted gene expression is expected. However, the highest overlap across all sets still occurred between colocalized eQTL and caQTL CSs not containing any ChromBPNet variant (13,253 shared incidences), while 27,026 caQTLs had no overlap with any other approach.

The molQTLs lacking ChromBPNet overlap is partly explained because ChromBPNet strictly models local chromatin accessibility at peaks, while most molQTLs occur outside of them (**Figures 1C, 4C**, in-peak caQTL category bars). C2 caQTLs were enriched in the caQTL-eQTL-ChromBPNet intersection relative to caQTL-eQTL-only loci (43.8% vs 21.5%; Fisher’s exact test OR = 2.85, 95% CI 2.66–3.06, p < 2.2×10⁻¹⁶; **Figure 4C**, in-peak caQTL category bars), indicating that ChromBPNet-overlapping caQTL-eQTL loci are biased toward the C2 caQTL class. Similarly, all three *caQTL-eQTL-ChromBPNet, eQTL-ChromBPNet,* and *caQTL-eQTL* showed similar patterns of cis-regulatory element (CRE) representation, with most occurrences falling within caPeaks defined as promoters and cis-CRE regions (±1Mb) (**Figure 4C**, caPeak annotation bars). These observations align with the previous observation (**Figures 1D-G**) and suggest that variants predictive of expression tend to span larger DNA regulatory regions.

We also observed that 65% of ChromBPNet significant variants are not found within any fine-mapped molQTL CSs (**Figure 4C**). When stratifying variant sets by minor allele frequency (MAF), ChromBPNet-specific variants show a higher proportion of lower allele frequencies (MAF > 0.05 & MAF < 0.1) than variants in other sets (**Figure 4B**, MAF bars). In addition, a large number of ChromBPNet-specific variants (47.5%) are located at chromatin accessible regions, lower density regions not defined as *peaks* in the caQTLs analysis, for which we used a more conservative peak-calling threshold (**Figure 4B**, caPeak annotation bars, See Methods). Indeed, variants privately predicted by ChromBPNet are located at chromatin accessible peaks with significantly lower accessibility amplitude per base-pair (*p*=8.32×10^52^) than ChromBPNet variants shared with molQTLs (**Figure 4D**).

Lastly, we compared the predicted ChromBPNet log(aFC) to the empirical caQTL beta for variants tested with both methods (**Figure 4E**). Significant variants in both methods showed a positive Pearson correlation (R^2^=0.451) in effect size and direction compared to method specific variants (caQTLs R^2^=0.034 and ChromBPNet R^2^=0.018). To further validate that these concordant effects reflect true allele-specific chromatin accessibility, we quantified allelic imbalance at heterozygous sites using phASERpop (**Figure S8A-B**). Allele-specific accessibility measurements showed high effect size correlation with both the empirical caQTL beta (Pearson’s R^2^=0.639) and the predicted ChromBPNet log(aFC) (Pearson’s R^2^=0.6), supporting the agreement between methods.

### Identifying instances of genetic variation reshaping transcription factor motif binding dynamics in CD4^+^ T cells

A key mechanism by which functional noncoding variants may influence chromatin accessibility is through altered TF binding ^40^. Using our CD4+ T cell-specific ChromBPNet models, we tested whether ChromBPNet-variants are associated with allelic differences in predicted accessibility contributions at TF motif-containing regions.

To establish a reference set of TF motif patterns captured by our CD4+ T cell ChromBPNet models, we applied TF-MoDISCo ^19,41^, which summarizes the sequence features learned by the model and recovers highly recurrent de novo motif patterns in this regulatory context. The discovered patterns largely matched known pioneers TF families within T cell lineage (e.g., NF-κB, IRF, AP-1, TCF/LEF, and ETS/RUNX) ^42–46^, yielding a final compendium of 20 TF families (**Table S2**). Most motifs were associated with positive predicted contributions to chromatin accessibility; in contrast, a de novo pattern resembling the PAX family (BH q_value_=0.00758) showed negative contributions, evident as an inverted contribution/importance profile (**Figure 5A**).

**Figure 5.**
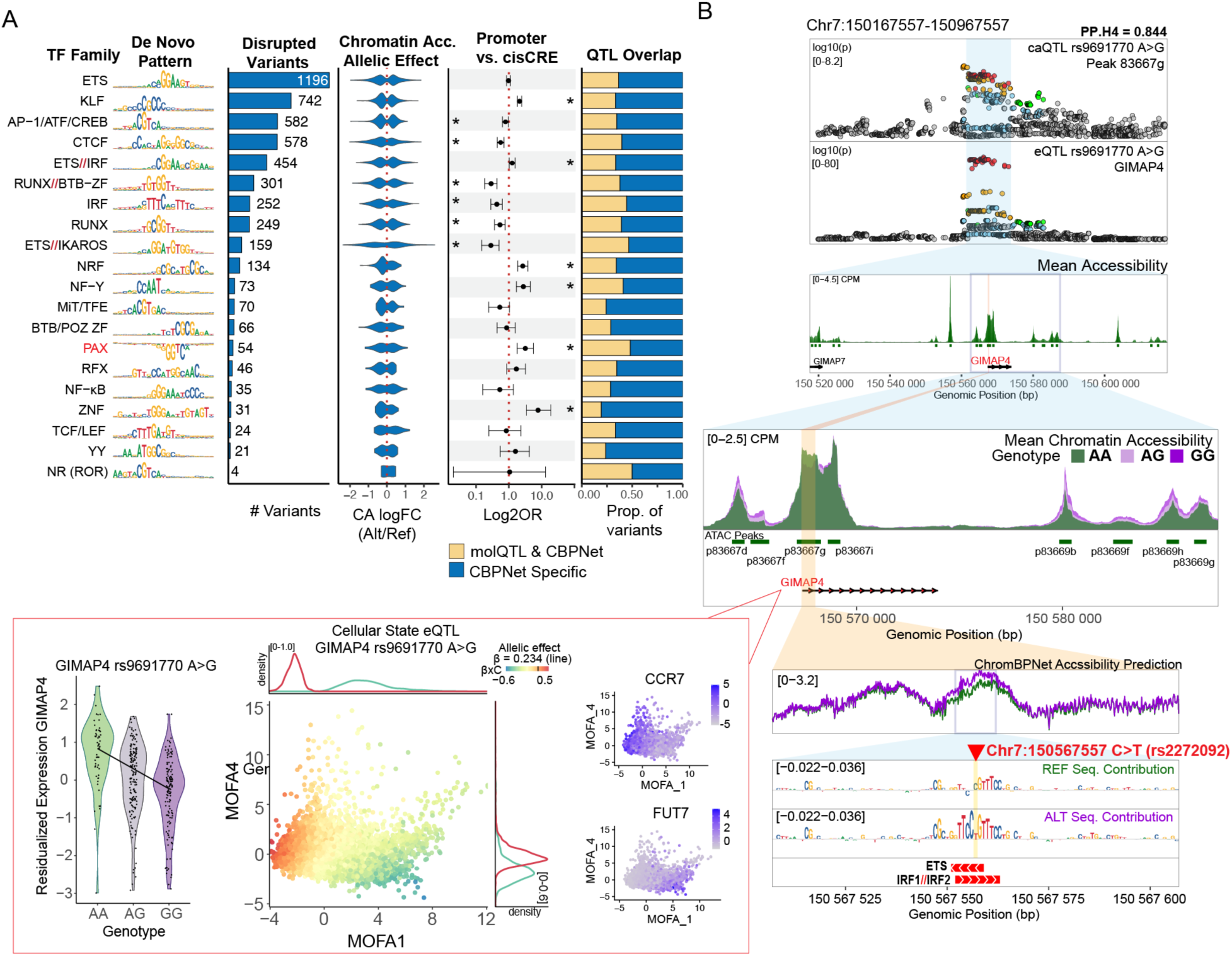
ChromBPNet implicates TF motif disruptions underlying colocalized molQTL effects. **(A)** Overview of TF motif family disruptions predicted for ChromBPNet-significant variants. Columns show: (1) motif family; (2) representative de novo motif pattern; (3) number of disrupted motif instances per family; (4) distribution of predicted allelic chromatin accessibility effects (allelic log fold-change, log(ALT/REF)) at disrupted instances; (5) enrichment of disrupted instances across genomic annotations (forest plots show odds ratios with 95% CIs; significance at FDR < 0.05, BH-corrected Fisher’s exact tests); and (6) composition of motif-disrupting variants that fall within molQTL credible sets versus ChromBPNet-only variants. **(B)** Example locus illustrating a ChromBPNet-nominated variant in an eQTL/caQTL-colocalized credible set with state-dependent regulation. Top: eQTL/caQTL colocalization at the locus. Middle: ATAC-seq coverage stratified by genotype (REF/REF, REF/ALT, ALT/ALT). Bottom: ChromBPNet allelic prediction for the candidate variant, with base-resolution contribution scores and allele-specific sequence logos (letter height reflects magnitude; direction reflects sign of contribution); (left) dynamic eQTL behavior for GIMAP4, violin plot shows the residualized expression of GIMAP4 across individuals with different genotypes, MOFA scatter plot shows the dynamic effect of the eQTL across the transcriptional states, and small MOFA scatter plots shows the expression of genes explaining the cellular state axis.

Next, we used Fi-NeMo to evaluate allele-specific TF motif disruption within 100-bp chromatin-accessible sequences centered on a variant. In total, 4,361 SNPs (38.1%) were predicted to directly perturb TF motif patterns, with both positive and negative predicted effects on local chromatin accessibility (**Figure 5A, S7E, Table S3**). Disruptions most frequently involved ETS, KLF, AP-1, CTCF, and RUNX family motifs (>500 hits each; **Figure 5A**), consistent with their broad occupancy across CD4+ T cell regulatory elements, reflected by the large number of TF-MoDISCo seqlets supporting these patterns (**Table S2**), and their established roles as core regulators of immune transcriptional programs ^42–44,47^.

SNP-disrupted TF motifs showed differential enrichment across regulatory contexts, with several TF families preferentially disrupted in promoter-associated peaks relative to distal cis-CREs. In particular, variants disrupting NRF, NF-Y, PAX, and ZNF family motifs were promoter-enriched, consistent with prior reports ^48–51^. Interestingly, SNP disruptions of RUNX, ETS, IRF, and AP-1 family motifs were more prevalent in distal cis-CRE peaks (**Figure 5A)**. We observed that 35.6% of the predicted TF disruptions were associated with ChromBPNet variants found within molQTLs CSs, whereas the remainder were TF-disruption-specific to ChromBPNet variants (**Figure 5A)**.

Figure 5B illustrates a colocalized eQTL/caQTL locus where the genetic signal (lead variant rs9691770; colocalization PP.H4 = 0.844) is associated with complex cell-state specific regulatory effects on GIMAP4. While the alternative allele (rs9691770-G) is associated with broadly increased chromatin accessibility across distal cis-regulatory elements surrounding GIMAP4 (chr7:150167557–150967557), at the promoter-proximal peak 63667g it correlates with decreased chromatin accessibility and decreased GIMAP4 expression, as depicted by the GIMAP4 expression violin plot and ATAC-seq coverage tracks.

Integrating ChromBPNet predictions at this molQTL CS, we identified a single significantly scored variant (rs2272092 C>T, with the T allele linked to rs9691770G, **Figure S6B**) predicted to increase local accessibility and enhance an overlapping ETS/IFR1/IRF2 motif instance proximal to GIMAP4 TSS, which are predicted by TF-MoDISCo^19,41^ to positively contribute to chromatin accessibility in our CD4⁺ T cell models (Figure 5A, **Table S2**).

Although this observation points to a direction mismatch between ChromBPNet predictions and caQTL data, it could also suggest a context-dependent repressive activity that this CRE might have on gene expression, as increased accessibility does not equal increased gene expression^52^. IRF1 and IRF2 recognize highly similar DNA motifs but exhibit antagonistic functions: IRF2 frequently acts as a competitive inhibitor of IRF1-driven transcriptional activation, dampening pro-inflammatory responses through competition at shared cis-elements ^53,54^. We hypothesize that the alternative allele may preferentially recruit IRF2 over IRF1 at this locus, resulting in increased local “openness” but reduced functional activity, coupling enhanced accessibility with transcriptional repression. This interpretation is further supported by the dynamic eQTL analysis showing strongest genotype-by-context interactions (G_βxC_) in naïve T cell states, where IRF2 expression and activity are known to be elevated. The GIMAP4-encoded GTPase regulates T cell survival, apoptosis, and effector functions, and is differentially expressed during Th1/Th2 polarization ^55–57^ consistent with context-dependent IRF-mediated regulation.

Together, these observations highlight that variant-centered, sequence-based predictions can capture local accessibility features while not necessarily recapitulating the net regulatory outcome at a locus, which may reflect state-specific TF occupancy/cofactor usage and multi-element cis-regulatory interactions beyond the modeled sequence window. This exemplifies how the integration of these approaches could reveal mechanistic subtleties potentially bypassed by using their method in isolation.

### Shared genetic signals between molQTLs and autoimmune diseases and traits

To provide regulatory context for GWAS disease variants, we colocalized (PP.H4>0.5) fine-mapped eQTLs and caQTLs from this study with a collection of 51 autoimmune diseases and traits derived from 97 GWAS (**Table S4)**. Across these immune traits, our molQTLs provided mechanistic support for a median of 33.3% of fine-mapped GWAS credible sets (Q1-Q3: 20.0-40.7%; Figure 6A**; Table S5**). Most of this signal was driven by caQTLs: caQTL-only colocalizations accounted for a median of 22.2% of GWAS credible sets per trait (Q1-Q3: 13.0-24.3%), and an additional 8.39% were supported by both eQTL and caQTL colocalization (Q1-Q3: 4.49-11.9%). In contrast, eQTL-only colocalizations contributed a smaller fraction overall (median 2.04%, Q1-Q3: 0-3.75%). Together, these results indicate that chromatin accessibility comprises an extensive, regulatory readout of genetic risk, capturing many loci with or without detectable expression colocalization, consistent with prior work ^15,58–61^. ChromBPNet variants overlapped a median of 4.57% of GWAS credible sets per trait (Q1-Q3: 0-7.48%), of which the majority of the signal was driven by QTL-shared variants (median 2.61%, Q1-Q3: 0-4.44%), while ChromBPNet-specific variants contributed a smaller fraction (median 0.69%, Q1-Q3: 0-2.98%, Figure 6A**, Table S6**).

**Figure 6.**
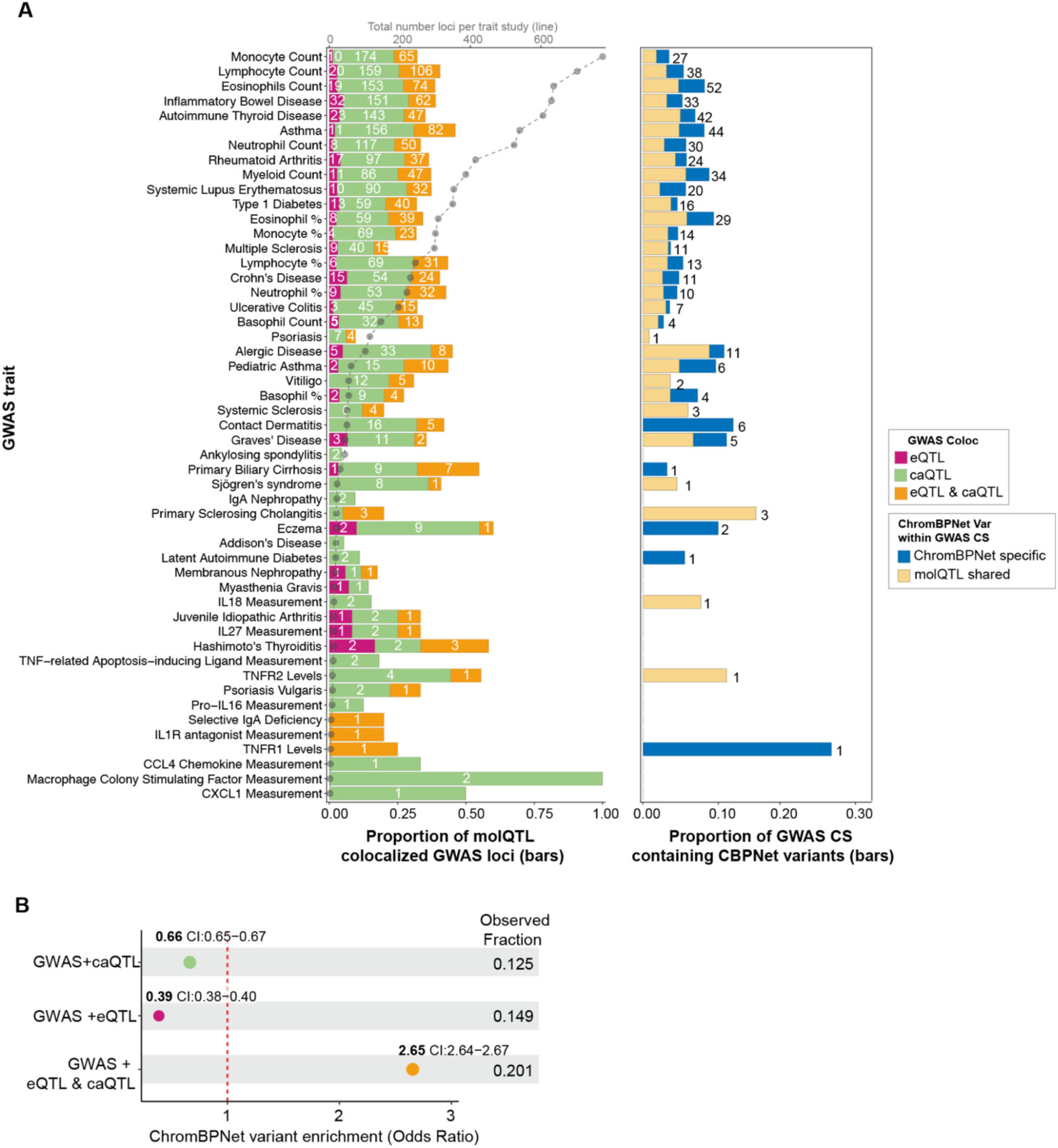
Colocalization of CD4⁺ T cell molQTLs and ChromBPNet variants with GWAS immune disease and trait loci. (**A**) **Left:** For each of 51 immune diseases/traits, barplots show the proportion of fine-mapped GWAS credible sets that colocalize with CD4⁺ T cell molQTL credible sets from this study, partitioned by colocalization class (eQTL-only, caQTL-only, or shared eQTL–caQTL); numbers on bars indicate counts of colocalized GWAS credible sets. An overlaid line plot from the top x-axis indicates the total number of fine-mapped GWAS credible sets per trait. **Right:** For each trait, barplots show the proportion of GWAS credible sets containing at least one ChromBPNet-prioritized variant (IPS empirical P < 0.05), stratified by whether the variant is shared with molQTL credible sets or is ChromBPNet-specific; numbers above bars indicate variant counts. (**B**) ChromBPNet hit enrichment in GWAS-colocalized molQTL credible sets. Odds ratios (95% CI) were estimated with a binomial GLM comparing each colocalization class (shared eQTL–caQTL, caQTL-only, eQTL-only) to non-colocalized credible sets; empirical one-sided P values were estimated from 100,000 permutations and reported as p < 1×10⁻⁵ when no permuted statistic equaled or exceeded the observed value.

To further understand which disease loci are captured by ChromBPNet, we tested the enrichment of ChromBPNet variants within molQTL colocalized GWAS loci (See Methods). We observed a strong GWAS enrichment of ChromBPNet hits in loci supported by both eQTL and caQTL colocalization (OR = 2.65, 95% CI: 2.64–2.67). In contrast, ChromBPNet hit fractions were depleted in caQTL-only loci (OR = 0.66, 95% CI: 0.655–0.673) and even more strongly depleted in eQTL-only loci (OR = 0.39, 95% CI: 0.380–0.396). All three comparisons were significant by permutation testing (*p* < 1×10⁻⁵) (Figure 6B). Together, these results suggest that ChromBPNet’s accessibility-based predictions are most concentrated at disease loci where chromatin and expression signals co-occur, consistent with a tighter mechanistic link between predicted local accessibility changes and downstream transcriptional effects.

## Discussion

In this study, we integrated empirical, regression-based molQTL mapping with cell-type-specific function-to-sequence chromatin accessibility predictive models (ChromBPNet) to orthogonally identify regulatory variants that modulate molecular phenotypes in CD4+ T lymphocytes. This framework enabled the detection of widespread cis-regulatory effects on gene expression and chromatin accessibility, the identification of context-dependent eQTLs, and prediction of variant-mediated TF binding disruption events. Together, these findings provide molecular context and mechanistic insight into genetic variants associated with 51 autoimmune diseases and traits identified by GWAS, shedding light on how genetic modulation of molecular programs within CD4+ T lymphocytes may shape inter-individual differences in immune response. However, we note that all analyses were performed from peripheral CD4+ T cells without additional stimuli.

Despite our single-cell data and dynamic eQTL mapping, our power to detect eQTLs in rare cell states or transient cellular subpopulations is limited, and these are unlikely to be well detected in our bulk ATAC-seq data. As a result, some context-dependent regulatory mechanisms, particularly those active only in specific activation states ^9,62,63^, may not be fully captured in the present study.

Our analyses indicate that caQTLs contribute to a substantial fraction of eQTLs and GWAS loci, consistent with prior studies highlighting the central role of chromatin-mediated regulation in complex traits ^15,58–61^. Causal inference analysis showed how expression can also regulate chromatin – rather than vice versa as is often assumed – with distinct regulatory architectures depending on the causal direction. A substantial fraction of caQTLs do not colocalize with eQTLs in our data, and some of these may represent context-dependent regulatory variants contributing to eQTL effects only under specific cellular or environmental conditions ^6^ or affect additional modes of genetic regulation that are not directly reflected in steady-state gene expression ^64,65^.

We used ChromBPNet and QTL mapping as two orthogonal approaches for variant prioritization. ChromBPNet and related sequence-based models are often applied to fine-mapped QTL or GWAS loci to prioritize likely causal variants ^16,66,67^, while our genome-wide analysis in conjunction with well-powered molQTL data allowed evaluation of their broader potential, which has remained less well characterized. Despite technical differences that complicate comparisons of these fundamentally different approaches, we observed that over half of the ChromBPNet variants were also supported by molQTL data with highly correlated effect sizes, and the remaining ones were enriched for low frequencies, likely representing true regulatory variants missed by molQTL analysis. The ability of sequence-based prediction to predict regulatory effects at nucleotide level rather than in haplotypes enabled easier inference of possible TF binding mechanisms. However, ChromBPNet detected only a subset of the tens of thousands of regulatory variants empirically detected by molQTL mapping, and less than 5% of GWAS loci. While this gap might be partially filled by other methods with scope to predict variant effects beyond open chromatin peaks ^17–19^, this may come at a cost in mechanistic interpretability.

Overall, this study underscores the importance of integrating multiple layers of information to discover and interpret genetic regulatory effects and their effects on disease. Multiple molecular phenotypes, integration with functional annotation data resources, and advanced analysis methods from variant effect prediction to causal inference allowed us to not only build catalogues of regulatory variants but also obtain insights into their mechanisms of action. Our results provide a quantitative framework for understanding the complementary strengths and limitations of empirical association-based and sequence-based prediction approaches, demonstrating the future potential of both methods.

## Data Availability

CD4T cell eQTL summary statistics, conditionally independent variants, and log bayes factors from SuSiE finemap are available at Zenodo (https://doi.org/10.5281/zenodo.18261455), as are the CD4T cell caQTL summary statistics, conditionally independent variants, and log bayes factors (https://doi.org/10.5281/zenodo.18317807).

Other data for this project, including ChromBPNet models, are available at Zenodo (https://doi.org/10.5281/zenodo.18394129)

https://doi.org/10.5281/zenodo.18261455

## Resource availability

CD4T cell eQTL summary statistics, conditionally independent variants, and log bayes factors from SuSiE finemap are available at Zenodo (https://doi.org/10.5281/zenodo.18261455), as are the CD4T cell caQTL summary statistics, conditionally independent variants, and log bayes factors (https://doi.org/10.5281/zenodo.18317807). ChromBPNet models and other data are available at Zenodo (https://doi.org/10.5281/zenodo.18394129). Scripts used to generate analysis and figures are available at https://github.com/marlmatos/cd4t-qtl-map

## Acknowledgments

This project was funded by R01AG057422 to TL and JMG from the National Institute on Aging (National Institutes of Health).

## Author contributions

M.R.M: NGS library preprocessing, conceptualization, data curation, formal analysis, investigation, visualization, methodology, writing original draft; writing review and editing

S.G: performed the data analysis molQTL and GWAS colocalizations, writing review and editing

S.B: data analysis and mediation analysis

M.R.M, K.L.P, R.D.T, J.S, T.V.T, MZ: Processed all blood samples

M.Z: Sample collection and NGS experimental design

D.R: scRNAseq library preparations

D.S, N.A, O.A: GWAS study collection and fine-mapping

A.G, M.I, S.R: Genotyping QC methodology support

S.G, S.R.O, K.R, S.M: Cohort Acquisition

T.L*: Conceptualization; supervision; funding acquisition; methodology; writing

J.M.G*: Conceptualization; supervision; funding acquisition; methodology; project administration

## Declaration of interests

T.L. is an advisor to Variant Bio with equity.

## Methods

### Cohort description

Individuals were recruited as part of the Albert Einstein College of Medicine Institute of Aging Research’s LifeLong and Logevity cohorts. The inclusion criteria were limited to healthy, self-identified Ashkenazi Jewish individuals, defined as having four Ashkenazi Jewish grandparents. After genotyping and data quality filtering, the final cohort in this study comprised 362 individuals (**Figure S1B**). Since the cohort was derived from an aging study, the age ranges are 20-41 years (median 27 years) and 66-100 years (median 78 years) for young and old, respectively.

### Sample collection and CD4+ T cell isolation

Peripheral blood was collected in 8 mL BD Vacutainer tubes (BD Biosciences) and transported to the processing site on ice. CD4⁺ T lymphocytes were isolated on the same day, with a median interval of 2.5 hours between phlebotomy and cell isolation. Cells were purified using the EasySep™ Human CD4⁺ T Cell Isolation Kit (STEMCELL Technologies, Cat# 17952) according to the manufacturer’s instructions. Purified CD4⁺ T cells were cryopreserved in CryoStor® CS10 (STEMCELL Technologies, Cat# 100-1061) at a density of 5 × 10⁵ cells/mL and stored in 2.0 mL cryogenic vials (Corning, Cat# 4435112) at −80 °C until downstream analyses.

### DNA sample preprocessing and genotyping

Samples processed for DNA sequencing were randomized across batches using the OSAT R package ^68^, accounting for covariates including age, sex, season, and technician (**Figure S1C**). For each batch, cryopreserved CD4⁺ T cells were thawed in a 37 °C water bath, and genomic DNA (gDNA) was extracted using the Quick-DNA Microprep Plus Kit (Zymo Research, Cat# D4074) according to the manufacturer’s protocol. DNA quality and concentration were assessed with a NanoDrop™ 1000 spectrophotometer (Thermo Fisher Scientific). Library preparation and low-pass whole-genome sequencing (target depth ∼1x) were performed by Gencove using their human low-pass GRCh38 v3.1 pipeline ^69^. Genotype imputation was carried out within this pipeline using an adapted version of GLIMPSE2 ^70^, with reference to the 1000 Genomes Project 30x panel ^71^. Following imputation, additional variant-level quality control was performed (**Figure S2A**). Using bedtools ^72^, variants with low genotype probabilities (*max[GP] < 0.9*) were excluded, while those with missingness <1% and minor allele frequency (MAF) >5% were retained. All subsequent quality control steps were conducted in PLINK v2.00a2.3 ^73^. Variants deviating from Hardy-Weinberg equilibrium were removed (*--hwe 1e-6*). At the sample level, duplicates and related individuals were excluded using KING kinship estimates (*--king-cutoff 0.0884*). To identify population outliers and derive genotype covariates, we LD-pruned autosomal variants (*--indep-pairwise 50 5 0.2*) and computed principal components (*--pca 50*, **Figure S9A**).

Population stratification was assessed using the 1000 Genomes Project ^74^ as a reference: reference genotypes were filtered with the same QC thresholds (*missingness 0.01, --hwe 1e-6, --indep-pairwise 50 5 0.2*) and merged with the study cohort before principal component analysis (*--pca*) ((**Figure S9B**). After genotyping quality control, 7.82 million variants and 377 genotyped samples remained available for association testing (**Figure S2A**).

### Sample Pre-processing for scRNA-seq and bulk ATAC-seq

For both scRNA-seq and ATAC-seq library preparation, samples were randomized and assigned to batches before thawing, resulting in 34 batches of 10-12 individuals each (**Figure S1E**). For each biological sample, a single vial of cryopreserved CD4⁺ T cells was thawed and split to generate both scRNA-seq and ATAC-seq libraries, ensuring that the two assays originated from the same biological replicate and thereby minimizing technical variability. Preprocessing prior to library preparation proceeded as follows. Cryopreserved CD4⁺ T cells were thawed in a 37 °C water bath until a small ice crystal remained. Cells were conditioned with lukewarm stabilizing buffer (RPMI supplemented with 10% heat-inactivated FBS) to neutralize the cryopreservation medium before transfer to 50 mL conical tubes (Corning Cat# 4435112). Cells were gradually reconstituted by sequentially doubling the stabilizing buffer volume (1 mL, 2 mL, 4 mL, 8 mL, up to 32 mL total). Tubes were gently inverted to mix and centrifuged at 400 rcf for 5 min at 4 °C. Cell pellets were resuspended in 2 mL Eppendorf Protein LoBind tubes (Cat# 022431102), and the centrifugation/washing step was repeated twice more with 1 mL of conditioning buffer each time to ensure complete removal of cryopreservation medium. CD4⁺ T cells were finally resuspended at a density of 1,250–2,500 cells/µL. We counted and assessed viability using propidium iodide staining on a Cellometer Auto 2000 (Nexcelom Biosciences) and proceeded only with cell suspensions containing greater than 85% viable cells.

### scRNA-seq library preparation and sequencing

We prepared 34 scRNA-seq dual-indexed pooled libraries (Chromium Next GEM Single Cell 3′ Kit v3.1 with Dual Index Kit TT Set A, 10x Genomics, PN-1000215) according to the manufacturer’s protocol (**Figure S1D-E)**. Library size and quality were assessed using a Fragment Analyzer (Agilent, SM101493-5200), and concentrations were determined with the KAPA Library Quantification Kit (Roche, Cat# 07960140001).

For sequencing, all dual-indexed libraries were pooled at equimolar concentrations and distributed across four S4 (16-lane) Illumina flow cells (**Figure S1D**). Sequencing was performed on the Illumina NovaSeq 6000 platform at the New York Genome Center using a paired-end 150 bp configuration (28/90/10/10 cycles). Libraries were sequenced across four NovaSeq S4 flow cells to a median depth of 3.19×10^8^ Read 2 (cDNA) reads per library (IQR: 2.81×10^8^-3.62×10^8^; totals summed across lanes from MultiQC/FastQC).

### scRNA-seq transcript quantification and library preprocessing

Dual-indexed, demultiplexed scRNA-seq library reads were aligned to the human reference genome GRCh38.p14. Alignment and gene quantification were performed with STARsolo ( -*-soloType CB_UMI_Simple, --soloFeatures Gene, --soloUMIlen 12*) and cell calling via *--soloCellFilter EmptyDrops_CR (*expected cells = 25,000; additional parameters: 0.99 10 45000 90000 500 0.01 20000 0.01 10000, STAR, v2.8.11b) ^75^. To enable allele-specific expression (ASE)–aware alignment, STAR was run with WASP tagging ^75,76^ (-*-waspOutputMode SAMtag)and supplied a cohort variant VCF via* --varVCFfile (MAF > 0.05, See Methods: *Sample Preprocessing and Genotyping*), with WASP-related BAM tags included in the output (--outSAMattributes NH HI AS NM MD vA vG vW). STARsolo count matrices were recalculated after filtering BAMs to autosomes and WASP-passing reads (vW = 1), using STARsolo gene/barcode/UMI tags (GX, CB, UB), so downstream analyses used only WASP-filtered reads.

Pooled libraries were demultiplexed by donor genotype using SouporCell (k = 12, v2.5) ^77^. Cluster identities were validated by Pearson correlation to matched WGS genotypes using Demuxafy ^78^. SouporCell-labeled singlets were retained; doublets and unassigned barcodes were excluded.

Quality control and preprocessing of singlet scRNA-seq data were performed with Seurat v5.67. For each library, cells were excluded if their transcript or gene counts deviated by more than two standard deviations from the library mean, or if >15% of reads mapped to mitochondrial DNA. For duplicated samples, we kept the instance with the highest cell count. In addition, we excluded SouporCell clusters that could not be confidently mapped to a sample in the metadata. The final dataset contained a median of 1,310 cells per individual (**Figure S10A**). Cells were annotated using Azimuth’s reference-based mapping using Azimuth’s PBMC reference ^79,80^. Lastly, we retained only cells with an Azimuth-predicted CD4⁺ T cell score > 0.8 (**Figure S10B**).

### Bulk ATAC-seq library preparation and sequencing

For each batch of 12 (See Methods: *Sample Pre-processing for scRNA-seq and bulk ATAC-seq*), nuclei were isolated using the Omni-ATAC protocol ^81^ with minor modifications. Aliquots of 70,000 viable CD4⁺ T cells were pelleted at 400 RCF for 5 min at 4 °C in a fixed-angle tabletop centrifuge (Eppendorf 5420). Pellets were gently resuspended in 50 µL of cold ATAC-seq lysis buffer (RSB1: 0.1% NP-40, 0.1% Tween-20, 0.01% digitonin) for 3 min, and the reaction was quenched with 1 mL of cold ATAC resuspension buffer (RSB2: 0.1% Tween-20 without NP-40 or digitonin) to reduce mitochondrial contamination. Nuclei were pelleted at 400 RCF for 10 min at 4 °C, the supernatant removed, and the nuclei pellet resuspended in 50 µL of transposition reaction mix (25 µL 2× TD buffer, 2.5 µL Tn5 transposase [100 nM final], 16.5 µL PBS, 0.5 µL 1% digitonin, 0.5 µL 10% Tween-20, 5 µL nuclease-free water). Transposition was performed at 37 °C for 30 min in a pre-heated thermocycler. Reactions were cleaned using the Zymo DNA Clean & Concentrator-5 Kit (Zymo Research, Cat# D4014), and DNA was eluted in 11 µL of elution buffer, yielding ∼10 µL of purified tagmented DNA for library amplification. Tagmented DNA was amplified in a 50 µL PCR reaction containing 25 µL NEBNext High-Fidelity 2× PCR Master Mix (New England Biolabs, Cat# M0541S), 10 µL unique dual indexes (Illumina, Cat# 20027213–15), 10 µL purified tagmented DNA, and 5 µL nuclease-free water. Thermocycling conditions were: 1 cycle of 5 min at 72 °C and 20 sec at 98 °C; 10 cycles of 10 sec at 98 °C, 30 sec at 63 °C, and 1 min at 72 °C. Amplified libraries were purified using a double-sided AMPure XP bead cleanup (Beckman Coulter, Cat# A63881) to remove both small (<100 bp) and large (>1,000 bp) fragments. Final library quality was assessed using the Qubit 1X dsDNA HS Assay Kit (Invitrogen, Cat# Q33230) on the Qubit4 fluorometer (Invitrogen, Cat# Q33238), and fragment size distribution was verified with the Bioanalyzer High Sensitivity DNA Kit (Agilent, Cat# 5067-4626).

ATAC-seq libraries were pooled at equimolar concentration across 8 sequencing batches (two NovaSeq S4 flow cells, **Figure S1D**) and sequenced on the Illumina NovaSeq600 at the New York Genome Center using a paired-end 150 bp configuration (151/151/10/10 cycles) with a target read depth of 50 million reads per library.

### Bulk ATAC-seq transcript quantification and preprocessing

We used a modified version of the nf-core/atacseq pipeline (based on release v2.1.2, Zenodo DOI: 10.5281/zenodo.2634132, ^82,83^) to preprocess ATAC-seq data. Raw sequencing reads were aligned to the human reference genome (assembly GRCh38.p14) using STAR v2.8.11b with WASP flags to account for allele-specific expression, as mentioned in scRNAseq preprocessing (see Methods). Aligned reads were filtered (samtools v1.18) to remove mitochondrial reads, unmapped reads, mate unmapped reads, secondary alignments, duplicate reads, reads failing quality checks, reads with incorrect variation flags, and sex chromosome reads (chrX, chrY) (-F 0×004 -F 0×0008 -f 0×001 -F 0×100 -F 0×0400 -F 0×0200 -q 1). Only properly paired reads with a minimum mapping quality of 1 and WASP flag vW=1 were retained. Additional filtering with bamtools retained only reads with insert sizes between -2000 and 2000 (**Figure S11A**), no more than 4 mismatches (NM tag ≤ 4), and no soft-clipped bases in their CIGAR strings. To correct for Tn5 transposase bias, alignments were shifted using deepTools alignmentSieve with the --ATACshift parameter. We confirmed expected enrichment of ATAC-seq signal at transcriptional landmarks by profiling signal around TSS and TES using deepTools (**Figure S11B**). For peak calling, all individuals’ processed ATAC alignments were merged, and peaks were called with MACS3 (-f BAMPE --nomodel -B --min-length 180 --max-gap 1000 -q 0.01 --nolambda --keep-dup all --SPMR). Individual-level transcript quantification was performed using featureCounts (subread) with a minimum fraction of overlapping bases of 0.2 (--fracOverlap).

### cis-eQTL mapping

For pan CD4⁺ T cell expression QTL mapping, we generated pseudobulk CD4⁺ T expression profiles for each donor by aggregating single-cell SCTransform-normalized counts using Seurat’s *AverageExpression* function ^84,85^. The resulting pseudobulk gene expression matrix was filtered to retain genes with less than 90% zeros across samples, and log(x+1) transformed and scaled the gene counts (zero mean, unit variance), following the approach described by Xue et al. (2024) ^86^ (https://github.com/powellgenomicslab/sc-veQTL). cis-eQTL were mapped with TensorQTL’s ^87^ using the *cis.map_nominal* and *cis.map_cis* functions (maf_threshold = 0.05, nperm=1000, fdr=0.05), while controlling for covariates including age, sex, number of cells, sequencing depth, batch, 11 genotype PCs, and 12 expression PCs (the latter defined by the elbow method using the PCAforQTL R package ^88^. To identify cis-eQTLs, we tested all genes with a TSS within a ±1Mb window of each variant. Multiple testing correction was performed using the Benjamini-Hochberg procedure to control the false discovery rate (FDR) at 5%.

### cis-caQTL mapping

Chromatin accessibility QTLs were mapped similarly to eQTLs (see Methods). For the phenotypes, the individual-level raw bulk ATAC-seq accessibility matrix was transformed to counts per million (CPM) using EdgeR and filtered to retain peaks with CPM ≥ 2 in at least 10% of samples. To reduce technical variability while preserving biology, we calculated a binned GC content for each peak given the reference sequence GRCh38 and applied a sample level GC-aware smooth quantile normalization using the qsmoothGC package ^89^. The resulting CPM matrix was filtered for missingness, log-transformed, and scaled as previously described. For the caQTL linear regression, we limited discovery to peak summits within a ±1Mb window from the variant tested, including age, sex, 23 ATAC PCs, and 15 genotype PCs as covariates.

### eQTL and caQTL statistical fine-mapping

For eQTLs, fine-mapping regions were defined as all SNPs within ±1Mb of each conditionally independent lead SNP. Overlapping regions were iteratively merged until no regions shared common SNPs, resulting in 968 disjoint fine-mapping regions. This ensured that each significant SNP and all SNPs within 500 kb of it belonged to exactly one region. The final regions ranged in size from 1 Mb to 11.9 Mb (median = 1.1 Mb). The extended MHC region (chr6: 25–36 Mb) was excluded due to its complexity and common practice of separate analysis. Statistical fine-mapping was performed using Sum of Single Effects (SuSiE) regression, as implemented in the susieR R package ^90^. SuSiE uses a Bayesian variable selection approach to identify credible sets of causal variants while accounting for linkage disequilibrium. We applied SuSiE regression with summary statistics (susie_rss) using the following parameters: L = 10 maximum causal variants per region, coverage = 0.95 for 95% credible sets, prior variance = 50, and residual variance estimation enabled. For regions that failed to converge at 95% coverage, we relaxed the coverage threshold to 0.10 with increased maximum iterations (1,000). Only regions with at least 100 variants were included in fine-mapping. The resulting CSs had a median size of 7 SNPs (IQR = 25).

For caQTLs, fine-mapping regions were defined as all SNPs within ±250 kb of each conditionally independent lead SNP. As with eQTLs, overlapping regions were iteratively merged to yield non-overlapping regions, resulting in 922 disjoint fine-mapping regions. Only variants with permutation-adjusted p-values < 0.01 were included in subsequent fine-mapping. Each significant SNP and all SNPs within 250 kb of it belonged to exactly one region. Region sizes ranged from 1 kb to 24.6 Mb (median = 1.2 Mb). The extended MHC region (chr6: 25–36 Mb) was excluded for the abovementioned reasons. Statistical fine-mapping was again performed using SuSiE regression (susieR) using the same parameters as described above, yielding CSs with a median size of 5 SNPs (IQR = 17).

### Colocalization between eQTLs and caQTLs

Colocalization analysis was performed to test whether eQTL and caQTL signals share causal variants. We used the coloc package, which implements an approximate Bayesian framework to estimate posterior probabilities for five hypotheses: H0 (no association), H1 (association with trait 1 only), H2 (association with trait 2 only), H3 (two independent causal variants), and H4 (shared causal variant). For eQTL-caQTL colocalization (eQTL-caQTL pairs), we used the coloc.bf_bf() method, which directly utilizes log Bayes factors (lbfs) from SuSiE fine-mapping outputs. Only credible sets identified by SuSiE were included in the colocalization analysis. Colocalization was performed for all gene-peak pairs within overlapping genomic regions containing at least 200 shared variants. We considered colocalization significant when the posterior probability of a shared causal variant (PP.H4) exceeded 0.5. For variants to be included in downstream interpretation, we additionally required: caQTL and eQTL nominal p-value < 0.001, and posterior inclusion probability > 0.01. In cases where SuSiE failed to converge for either QTL signal, we performed single-variant colocalization using the first component of the log Bayes factor matrix. Single-SNP posterior probabilities under H4 were calculated as exp(lbf₁ + lbf₂ - logsum(lbf₁ + lbf₂)), and region-level hypothesis posteriors were computed using the combine.abf() function with default prior probabilities (p1 = 1×10⁻⁴, p2 = 1×10⁻⁴, p12 = 1×10⁻⁵).

### Allele specific quantification

Allele-specific chromatin accessibility was quantified with phASER ^91^ by integrating donor-matched ATAC-seq alignments with phased genotypes. Cohort genotypes were statistically phased with SHAPEIT4 ^92^ using the phase 2 1000 Genomes GRCh38 reference haplotype panel (https://ftp.1000genomes.ebi.ac.uk/vol1/ftp/technical/reference/phase2_reference_assembly_sequence/). For each donor, phASER was run on paired-end ATAC-seq BAM files to estimate haplotype-resolved read support at heterozygous sites, applying read-quality thresholds (MAPQ ≥ 30, base quality ≥ 10). Using the phASER *Gene AE* function*, f*eature-level haplotypic counts were summarized by quantifying reads overlapping each haplotype within each genomic feature (e.g., caQTL peak coordinates or ChromBPNet ATAC-seq peak sets). Finally, we used *phASER-POP* to align haplotype labels across donors using the multi-sample phased genotypes together with feature-variant pairing and sample mapping information, yielding population-level allelic counts with consistent allelic orientation for downstream analyses.

### caQTL categories

To better resolve putative causal mechanisms, we classified fine-mapped caQTL variants by their position relative to the associated chromatin accessibility peak (caPeak). Because each caQTL credible set contains one or more variants, we assigned categories at the variant level and then derived a single CS-level label using a hierarchical rule. At the variant level, each variant was assigned to one of three classes: C1, overlapping its associated caPeak; C2, overlapping a chromatin accessibility peak other than its associated caPeak; or C3, not overlapping any chromatin accessibility peak. At the CS level, CSs were annotated hierarchically: in_caPeak if the CS contained ≥1 C1 variant; in_other_Peak if no variants were C1 but ≥1 variant was C2; and no_Peak_overlap if all variants were C3.

### Integrative characterization of fine-mapped molQTLs

#### molQTL relative distance to eGenes/caPeaks

We quantified the genomic proximity between molQTL signals and their linked regulatory features using fine-mapped credible sets (CS) as the unit of analysis. For each CS, we calculated the mean absolute distance between all variants in the CS and the associated feature. For eQTLs, distances were computed to the linked eGene transcription start site (TSS) using GENCODE v44 (hg38) gene annotations (Accession ENCFF651QPF). For caQTLs, distances were computed to the summit of the linked ATAC-seq caPeak from the union peak set (see Methods: Bulk ATAC-seq transcript quantification and preprocessing). Differences in distance distributions were assessed using non-parametric rank-based tests on log-transformed distances (log2[distance to caPeak + 1]). Specifically, we compared eQTL vs. caQTL distance distributions using a Wilcoxon rank-sum test (caQTL categories collapsed), and tested for differences among caQTL overlap categories using Kruskal–Wallis with post-hoc pairwise Wilcoxon tests; p-values for pairwise category comparisons were adjusted for multiple testing using the Benjamini-Hochberg (BH) procedure.

#### Number of eGenes/caPeak per locus

To quantify target multiplicity per locus for eQTLs and caQTLs, we used fine-mapped credible sets (CS) as the unit of genetic signal. We summarized the distribution of CS sizes (number of variants per CS; **Figure S4B**) and, to limit the influence of unusually large CS on locus span and overlap, retained CS in the bottom 75th percentile of this distribution for each QTL type. For each retained CS, we defined a genomic interval using the minimum and maximum variant positions and computed pairwise overlap between CS intervals using the Jaccard index (intersection/union). CS pairs with Jaccard ≥ 0.3 were grouped into loci, defined as connected components of the overlap graph. We then quantified locus-level “pleiotropy” by counting the number of unique targets associated with any CS in each locus (unique genes for eQTL loci; unique peaks/features for caQTL loci) and summarized these counts into target-multiplicity bins. Differences in bin proportions between eQTL and caQTL loci were assessed using a Pearson’s chi-square test of independence (comparing multiple proportions across bins), and controlled for multiple testing using BH correction.

#### Genomic region and chromatin-state annotations of fine-mapped credible sets

For eQTL and caQTL separately, CS size was defined as the number of distinct variants per CS, and we retained CS in the bottom 75th percentile to reduce the influence of unusually large CS (**Figure S4B)**. For genomic region annotation of eQTL variants, we used GENCODE v44 (hg38 primary assembly) and locateVariants, assigning each variant a single label using the priority coding > fiveUTR > threeUTR > spliceSite > promoter > intron > intergenic (fiveUTR/threeUTR optionally collapsed to UTR). Each CS was assigned a single genomic-region label by majority vote across its variants, breaking ties by the same priority. For chromatin state, we intersected unique variants with an ENCODE T-cell chromatin-state segmentation (ENCODE accession ENCFF651QPF) and collapsed raw states into six groups: Promoter, Enhancer_active, Enhancer_weak, Transcribed, Repressed_polycomb, ZNF_Het_Quies (mapping defined in code). CS-level chromatin state was assigned by majority vote across variants, with ties broken by group priority Promoter > Enhancer_active > Enhancer_weak > Transcribed > Repressed_polycomb > ZNF_Het_Quies. Differences in chromatin-state composition were tested using Pearson’s chi-square tests: eQTL vs caQTL_all (collapsed) and pairwise comparisons among caQTL categories, with per-state state-vs-rest 2×2 tests and BH correction across states.

#### Colocalization summary across molQTLs

We summarized molQTL colocalization using fine-mapped credible sets (CS) as the unit of association. For each CS, we used statistical colocalization to classify signals as shared between molQTLs versus type-specific based on posterior support for a common causal variant (PP.H4 > 0.5; see Colocalization Methods). We then reported the fraction of CS in each class for eQTLs and caQTLs, and for caQTLs further stratified these fractions by peak-overlap category (in_caPeak, in_other_Peak, no_Peak_overlap). Differences in the fraction of shared signals across caQTL categories were assessed using pairwise chi-square tests of independence (colocalized vs. not colocalized), with p-values adjusted across category pairs using BH correction (**Figure S4F**).

### Dynamic-eQTL mapping

Dynamic eQTL interactions with cellular state were tested using CellRegMap^23^. Single-cell preprocessing was performed in Scanpy (v1.10.3). After standard preprocessing (see *scRNA-seq Transcript Quantification and Library Preprocessing*), we applied stricter QC within each CD4⁺ T subtype, removing cells with outlier detected genes (5th–95th percentiles), high total counts (>99th percentile or >10,000), low log-counts (<1st percentile; <5th for cytotoxic cells), or high ribosomal fraction (>98th percentile) (**Figures S5A-C**). After filtering, a total of 372,838 (88.3%) cells were retained for downstream analysis (**Figures S5D**). Raw counts were CPM-normalized, log2-transformed, and batch-corrected with Harmony using 50 PCs.

Metacells were generated by clustering cells within each donor across Leiden resolutions 3.4, 4.0, 4.5, and 5.0 and aggregating counts per donor×cluster (**Figures S5E-F**). Cellular states for dynamic interaction testing were defined by reducing the metacell’s single cell matrix into latent factors using Multi-Omics Factor Analysis (MOFA) ^25,26^, as performed by the CellRegMap authors ^23^. MOFA models (MOFApy2 v0.7.1) were fit in parallel for each pseudocell matrix (one per Leiden resolution) using the 500 most variable genes and 50 PCs, with K = 5 factors (convergence_mode = “medium”). We carried forward the Leiden 3.4 pseudocell matrix for downstream testing based on variance explained and factor independence (**Figures S5G-H**), yielding 17,758 pseudocells (10–35 cells/pseudocell; 30–60 pseudocells/donor) (**Figures S5E-F**). To annotate cellular states defined by MOFA factors, we performed GO Biological Process enrichment for each factor using the genes with the largest absolute loadings (hypergeometric test; background: the 500 variable genes used to fit MOFA). P values were Benjamini-Hochberg adjusted (FDR < 0.05). Redundant terms were collapsed using Wang semantic similarity (cutoff = 0.7), retaining the most significant term (lowest adjusted p-value) per similarity cluster (**Figures S5I-J**).

For interaction testing, we utilized conditionally independent cis-eQTLs discovered in the previous section, retaining the top variant per eGene. Each cis-eQTL was tested across donor metacells for genotype-by-context effects using the K = 5 MOFA factors, adjusting for donor age and sex. Significant interactions were called at FDR < 0.05 and are referred to as dynamic eQTLs. For significant interactions, we extracted effect-size estimates for the persistent genetic effect (β_G_) and the genotype-by-context component (β_G×C_). Dynamic-eQTL visualizations were based on visualizations provided by the CellRegMap^23^ authors in their GitHub repository (https://github.com/annacuomo/CellRegMap_analyses)

### Mediation analysis

We used bidirectional mediation analysis to infer causal relationships between accessible chromatin regions and variation in gene expression. Genetic variants were used as exposure variables (E) in a causal inference framework, where chromatin accessibility and gene expression measurements were used as either the mediator (A) or outcome (B) depending upon the direction that was being tested.

Causal estimates were inferred between A and B using the Findr ^39^ package (version1.0.8) in Python (version 3.12.3), which uses likelihood ratio tests to infer causal relationships between pairs of continuous variables. Causal estimates for P(A->B) were obtained from a composite test, combining posterior probabilities of the secondary linkage test and the conditional impendence test from the Findr package. The secondary linkage test measures the association between the E and B, whereas the conditional impendence test assesses independence between E and B when conditioning on A.

When testing the causal effect of chromatin accessibility upon gene expression, chromatin accessibility trait loci (caQTLs) were used as exposure variables, chromatin accessibility measurements were the mediator and gene expression was the outcome. In the case of gene expression to chromatin accessibility, expression quantitative trait loci (eQTLs) acted as the exposure variables, gene expression was the mediator and chromatin accessibility the outcome.

Individual-level data were required inputs to Findr, and both gene expression and chromatin accessibility measurements were corrected for age and sex prior to analysis and underwent rank-based inverse normal transformation within the Findr package. For all previously described colocalization events, we tested both the colocalized gene and chromatin peak as mediators against all possible outcomes as exposures, as required for false discovery rate (FDR) distribution estimation. For each mediator, the lead variant (highest PIP) of the fine-mapped credible set for the mediator was used as the exposure variable. From all calculated posterior probabilities for P(A→B) we estimate a global FDR as:

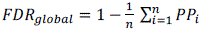

Different P(A→B) thresholds were implemented in a stepwise manner until the desired FDR global threshold was obtained. Using previously described fine-mapped information, it was then possible to map causal estimates for colocalized triads to the mediation results.

### Locus-level summarization of colocalized eQTLs and caQTLs

To avoid counting the same genetic signal multiple times, we treated each colocalized eQTL-caQTL credible-set pair (PP.H4 > 0.5) as a single signal and collapsed all associated variant-chromatin-gene triplets into one locus ID for downstream post-mediation analyses. Figure 3B summarizes locus collapsed mediation events.

### Feature enrichment across mediation models

To compare the chromatin-to-gene and gene-to-chromatin mediation models (Figure 3E-F), we tested whether peak-gene pairs assigned to each model were enriched for specific annotations. First, we quantified enrichment of peak position relative to the linked gene (promoter-proximal, intragenic, upstream, downstream) between models. Second, we tested enrichment of caQTL overlap categories between models. In both analyses, enrichment was assessed using logistic regression (binomial GLM with logit link) with model class as the predictor and annotation membership as a binary outcome, reporting odds ratios (OR) with Wald 95% confidence intervals and two-sided p-values. To account for non-independence of multiple triads within a locus, we used locus-clustered robust standard errors, and controlled for multiple testing across classes using Benjamini-Hochberg correction.

### ChromBPNet model for pan CD4⁺ T cells

Pan CD4⁺ T cell chromatin accessibility models were trained using ChromBPNet (github:kundajelab/chrombpnet:latest, v1.0, commit# aae277e)^16^ on eight samples with >100 million ATAC-seq reads each (**Figure S7B**). Reads from these samples were pooled, and peaks were re-called (hereon referred to as ChromBPNet peaks) with a looser threshold than that used in QTL ATAC-seq preprocessing (see Methods) using MACS2 (-f BAMPE --nomodel --extsize 183 -p 0.01 --call-summits). Lastly, peaks were filtered to exclude GRCh38 blacklisted regions (ENCODE accession ENCFF356LFX). To prepare for the model training, the peak regions were used to create non-peak genomic background regions with similar GC content to the peak regions. Both peak and non-peak regions were then partitioned into training, validation, and testing sets across five chromosome-based folds (see folds 0-4 in **Figure S7A**). We chose only fold-0 to train the *ChromBPNet Bias model*, which learnt the inherent sequence bias of the tn5 transposase used in the ATAC-seq library preparation. We trained multiple *bias models* using the GRCh38 reference genome and evaluated them using the performance metrics recommended by the authors, with the best-fit iteration retained for downstream analyses *(chrombpnet bias pipeline -b 0.7)*. The trained fold-0 *ChromBPNet Bias model* was used as input to train the *ChromBPNet bias-factorized model (chrombpnet pipeline),* following the procedures described by Pampari et al.^16^ (https://github.com/kundajelab/chrombpnet). Parallel Bias-Factorized *ChromBPNet models* for each fold 0-4 were trained (model characteristics summarized in **Figure S7C**) and subsequently used to calculate prediction and SHAP contribution count/profile scores for each fold. For downstream analyses, we averaged sequence predictions and SHAP contribution profiles across all folds.

### DeNovo TF motif discovery

To identify de novo TF motifs contributing to chromatin accessibility in CD4⁺ T cells, we applied TF-MoDISco ^41^ using the implementation provided in the ChromBPNet package (singularity image github:kundajelab/chrombpnet:latest, v1.0, commit# aae277e)^16^. We identified de novo motif patterns with TF-MoDISco (*modisco motifs -n 1000000 -w 400*) using averaged SHAP sequence contribution count scores as input. We retained only motif patterns supported by at least 40–50 seqlets and excluded those with low visual complexity or with >50% GC content. For motif interpretation, we merged a collection of position weight matrices (PWMs) from three reference databases JASPAR ^93^, HOCOMOCO ^94^, and CIS-BP ^95^. We supplied the motif PWMs to the TF-MoDISco report utility, which employs TOMTOM ^96^ to match de novo motifs to known TF motifs. Significant TF motif matches (q < 0.05) were annotated with the corresponding TF family. When multiple significant known motif matches were detected for a pattern, we assigned a shared TF-family label (e.g., ETS/IRF). Motifs that closely resembled a known motif but did not meet significance were provisionally annotated with the closest TF name and flagged as not significant, while low-complexity or unassignable non-significant motifs were excluded from downstream analyses.

### ChromBPNet variant scoring

To identify variants predicted to influence chromatin accessibility, we applied the trained CD4⁺ T cell *bias-factorized ChromBPNet model* ^16^ to score genotyped variants within our cohort. Given model limitations ^16^, the initial set of imputed variants (7 million variants, MAF>0.05) was filtered to retain SNPs located within ChromBPNet peak set. Using the *variant-scorer* helper scripts (https://github.com/kundajelab/variant-scorer, commit# 0e1e341)^16^, we predicted the sequence contributions of 159,908 SNPs (window=1000bp). Variant significance was evaluated using the Integrative Prioritization Score (IPS), which combines the Active Allele Quantile (AAQ), allelic log fold change (logFC), and Jensen–Shannon distance (JSD), as recommended by Pampari et al. (2025)^16^. Variants with an absolute IPS empirical p-value < 0.05 were prioritized as significant.

### Predicting TF motif disruption

To predict disrupted TF motifs, we employed the function *hitcaller* within (https://github.com/kundajelab/variant-scorer, commit# 0e1e341) which uses Fi-NeMo (https://github.com/austintwang/finemo_gpu, commit# 5c6f521), a tool that leverages ChromBPNet’s sequence contribution scores along with TF-motif patterns previously identified by TF-MoDISco to identify matching TF motif instances across the genome. To identify allele-specific TF-motif hits overlapping likely disrupted by the presence of a genetic variant, we supplied Fi-NeMo with variant contribution scores and their genomic locations. Motif hits were filtered to keep hits with a match_correlation > 0.8, hit_coefficient > 5, hit_importance>0.1.

### Characterizing ChromBPNet variants concordance with eQTL and caQTLs

We integrated ChromBPNet-predicted variants with fine-mapped caQTLs and eQTLs to compare predictive concordance among functional variant-prioritization methodologies. We defined ChromBPNet variant membership within fine-mapped molQTLs as a QTL credible set that contains at least one ChromBPNet variant. This membership approach resulted in seven distinct interception groups (caQTL only, caQTL+eQTL, chromBPNet only, chromBPNet+caQTL+eQTL, chromBPNet+caQTL,eQTL only, eQTL+chromBPNet) that quantified overlapping incidences between caQTLs, eQTLs, and ChromBPNet variants, rather than the number of variants shared. We characterized these mutually exclusive sets of variant incidences by quantifying the relative proportion of variants within caQTL context categories (See Methods *ca-QTLs Context Categories*), chromatin accessibility informed genomic regulatory regions (See Methods *Chromatin accessibility-informed regulatory element classification*), and minor allele frequencies binning for variants within these sets.

We further compared allelic effects for all ChromBPNet-scored variants against their corresponding empirical caQTL associations in both magnitude and direction. Because the sets of variants evaluated by each approach are not identical, owing to differences in peak definitions, variant types (SNPs versus indels), and genomic inclusion criteria, we first harmonized both datasets by restricting the analysis to SNPs present in both. We further limited the comparison to caQTL variants located *within* chromatin accessibility peaks (C1 and C2 caQTLs), as ChromBPNet predictions are confined to local regulatory effects. To assess concordance between methods, we compared ChromBPNet-predicted allelic effects on accessibility (log fold-change, ALT/REF) from the variant-scorer framework with caQTL effect sizes (β, ALT/REF) estimated using TensorQTL. For variants significant in both approaches, we used Pearson’s correlation coefficient to quantify agreement in the direction and magnitude of allelic effects.

### Chromatin accessibility-informed regulatory element classification

We defined regulatory elements based on a chromatin accessibility informed framework. We utilized chromatin accessibility peaks used in either, caQTL mapping (Figure 4C) or ChromBPNet peak sets (Figure 5A). Using Gencode v44, we annotated any chromatin peak overlapping a TSS as a *promoter* and those not overlapping as a *cis-CREs*.

### Quantifying Peak Coverage overlapping ChromBPNet variants

Read coverage for peak regions was quantified as previously described (featureCounts with --fracOverlap 0.2; see Methods, “*Bulk ATAC-seq transcript quantification and preprocessing*”), but here applied to the ChromBPNet input peak calls. Quantified peak regions were then intersected with ChromBPNet variants by genomic position, retaining only overlapping peaks. We next compared the distribution of chromatin accessibility coverage between ChromBPNet-specific variants and ChromBPNet variants found within any molQTL (by CSs membership as previously described), using a Student’s *t*-test (Figure 4D).

### Enrichment of variant disrupted TF motif families in promoter vs. cis-CREs

To test whether motif-disrupting variants for each TF family are enriched in promoters versus cis-CREs, we performed a Fisher’s exact test using **variants as the unit** (each variant counted at most once per family). The universe comprised all ChromBPNet-tested variants within the union ChromBPNet peak set, with peaks annotated as promoter or cis-CREs (see Methods), and each variant inheriting the label of its overlapping peak; for each family, we collapsed hits to unique variant×family pairs, built a 2×2 contingency table (hit vs non-hit by promoter vs cis-CREs) to estimate odds ratios (95% CI), applied BH correction across families, and excluded families with low hit counts.

### Data Visualization for Genetic Analysis

Genomic track plots (locus plots, ATAC-seq coverage, and ChromBPNet contribution score sequence logos) and TF motif PWM were generated with custom visualization code adapted and extended from Liu et al. 2025 ^97^

### QTL Colocalization with GWAS traits

We performed colocalization of our eQTLs and caQTLs to 88 independent GWAS for immune diseases and blood traits (**Table S4**). To enable comparison with GWAS summary statistics reported in GRCh37, fine-mapped QTL results initially generated in GRCh38 were converted to GRCh37 coordinates. Variant positions were lifted-over using bedtools intersect (v2.31.1) with dbSNP build 151 for both GRCh38 and GRCh37. Variants were matched across genome builds using rsID identifiers, and both forward and reverse allele orientations were tested to ensure proper allele alignment. Pairwise colocalization was performed using SuSiE COLOC ^98^ between each GWAS and each molQTL. GWAS were fine-mapped using in-sample or ancestry-matched linkage disequilibrium where possible. SuSiE COLOC ^98^ was conducted with default settings using log Bayes factors generated with SuSiE fine-map. Where fine-mapping did not converge or colocalization failed, COLOC under the single causal variant assumption was performed. To quantify the proportion of GWAS loci that colocalize with molQTLs in this study, we grouped GWAS studies by trait. We calculated both the total number of CSs and the number of CSs that colocalized with at least one molQTL. To quantify the proportion of GWAS CSs explained by ChromBPNet, we used a membership-based approach, as described previously, by counting the number of unique CSs that contained at least one ChromBPNet-scored variant relative to the total number of CSs per trait.

### Enrichment of ChromBPNet variants in GWAS-colocalized QTL credible sets

To test whether ChromBPNet-predicted regulatory variants are enriched in GWAS-colocalized molQTL loci, we classified fine-mapped molQTL credible sets (CSs) by GWAS colocalization status (shared eQTL–caQTL, caQTL-only, eQTL-only) and used non-colocalized CSs as the background, excluding variants in the extended MHC (chr6:25–34 Mb). For each CS, we defined “callable” variants as those scored by ChromBPNet and “hits” as callable variants passing our IPS significance threshold (p < 0.05) and summarized each CS by the number of hits ((h)) out of callable variants ((m)). Enrichment was estimated using a binomial generalized linear model comparing each colocalization class to the background (hit vs non-hit counts: (h) vs (m-h)), adjusting for chromosome and callable set size, and reported as an odds ratio (OR) with 95% confidence intervals. Statistical significance was assessed with an empirical null by simulating hit counts per CS from a binomial distribution using chromosome-specific background hit rates estimated from non-colocalized CSs (10^5^ permutations) and computing a one-sided permutation p-value based on the mean hit fraction across CSs in each colocalization class.

## Notes

### Competing Interest Statement

T.L. is an advisor to Variant Bio with equity.
The other authors declare no competing interests.

### Author Declarations

The Institutional Review Board of the Albert Einstein College of Medicine gave ethical approval for this work

