## Supplemental Figures for "Integrating multi-omic QTLs and predictive models reveals regulatory architectures at immune related GWAS loci in CD4+ T cells"

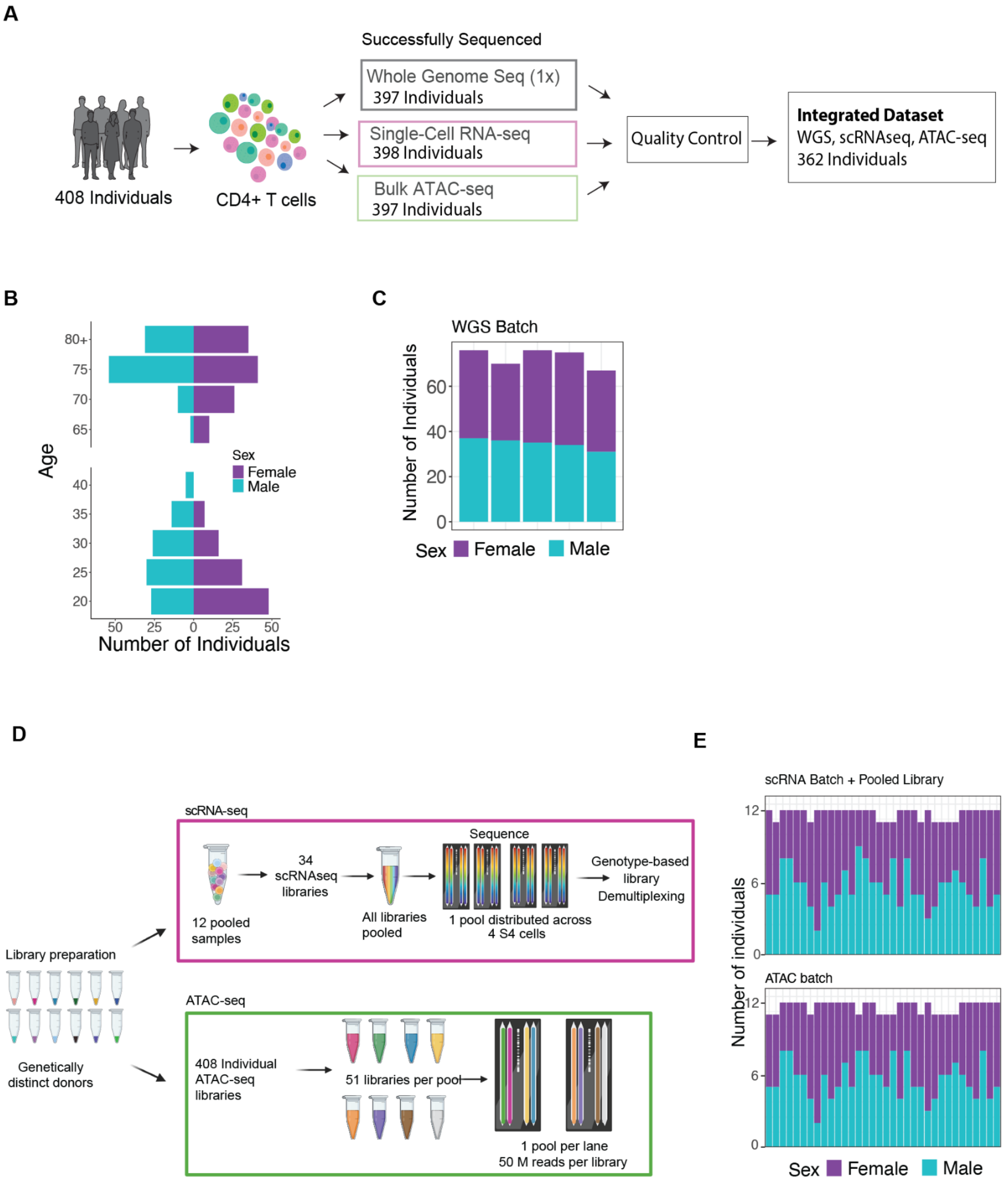

**Figure S1. Study design, cohort characteristics, and sequencing/QC summaries.** (**A**) Overview of the study design and multiomic data generation across donors. (**B**) Cohort age distribution stratified by sex (pyramid plot). (**C**) Batch distribution of low-pass whole-genome sequencing libraries, colored by sex. (**D**) Schematic of the tandem batching and sequencing strategy for scRNA-seq and ATAC-seq libraries. (**E**) Batch randomization and composition of donors across scRNA-seq and ATAC-seq batches, colored by sex.

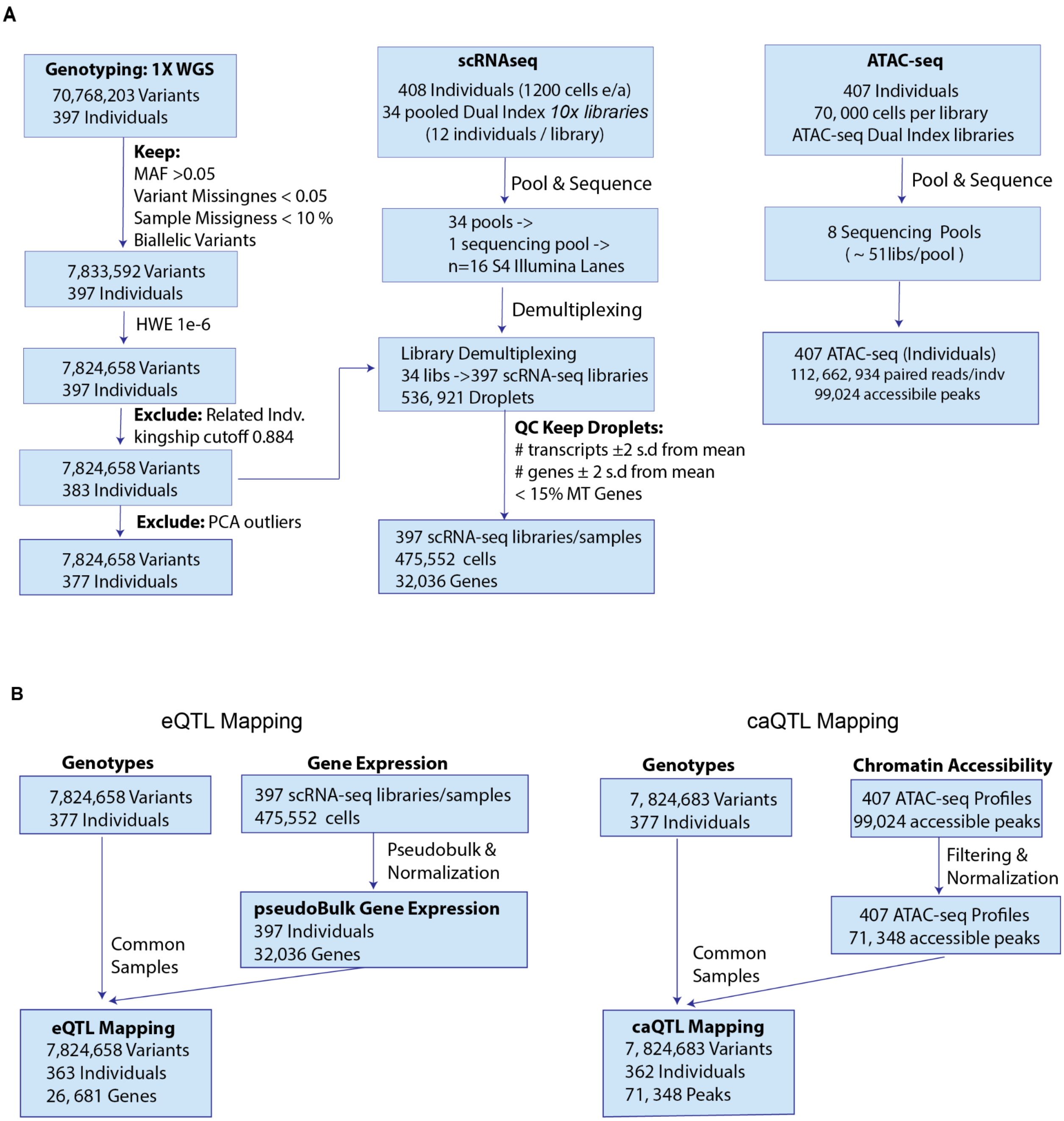

**Figure S2. Quality control for sequencing datasets.** (**A**) Flowcharts summarizing the QC and preprocessing workflows for each data modality: (*left)* low-pass whole-genome sequencing; (*middle)* scRNA-seq; (*right)* ATAC-seq. (**B**) Flowcharts summarizing the inputs used for molQTL discovery, including the number of samples, variants, genes, and peaks retained after QC and filtering (left, eQTL; right, caQTL).

**Figure S3**

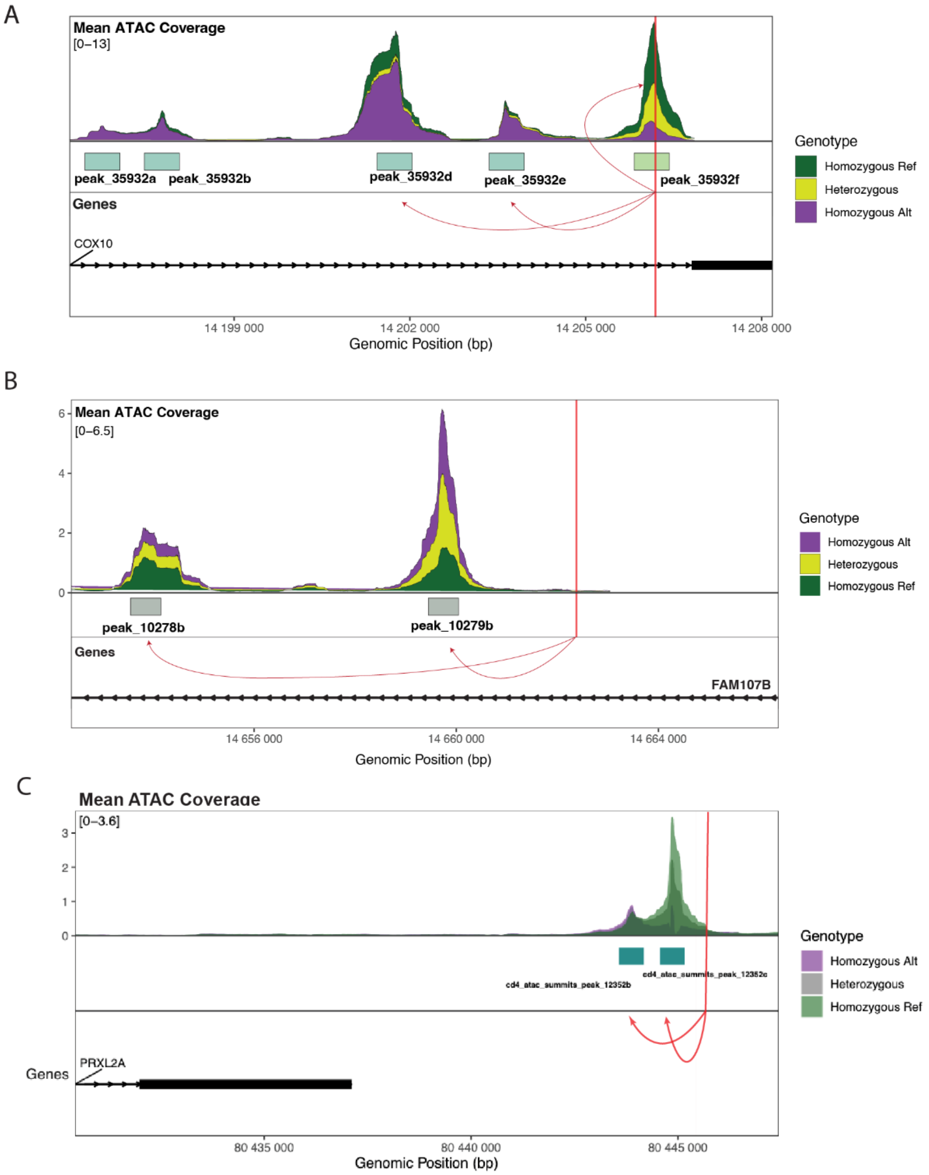

**Figure S3. Examples of caQTL categories.** ATAC-seq coverage tracks stratified by caQTL genotype (individuals grouped by genotype), illustrating representative caQTL positional categories relative to called chromatin peaks. Arrows depict variant-peak associations. (**A**) Examples of in-peak caQTLs (C1) and in-other-peak caQTLs (C2). (**B**) Example of a C3 caQTL in which the lead variant does not overlap any called peak. (**C**) Additional C3 example where the lead variant lies proximal to, but outside, the statistically defined peak boundaries.

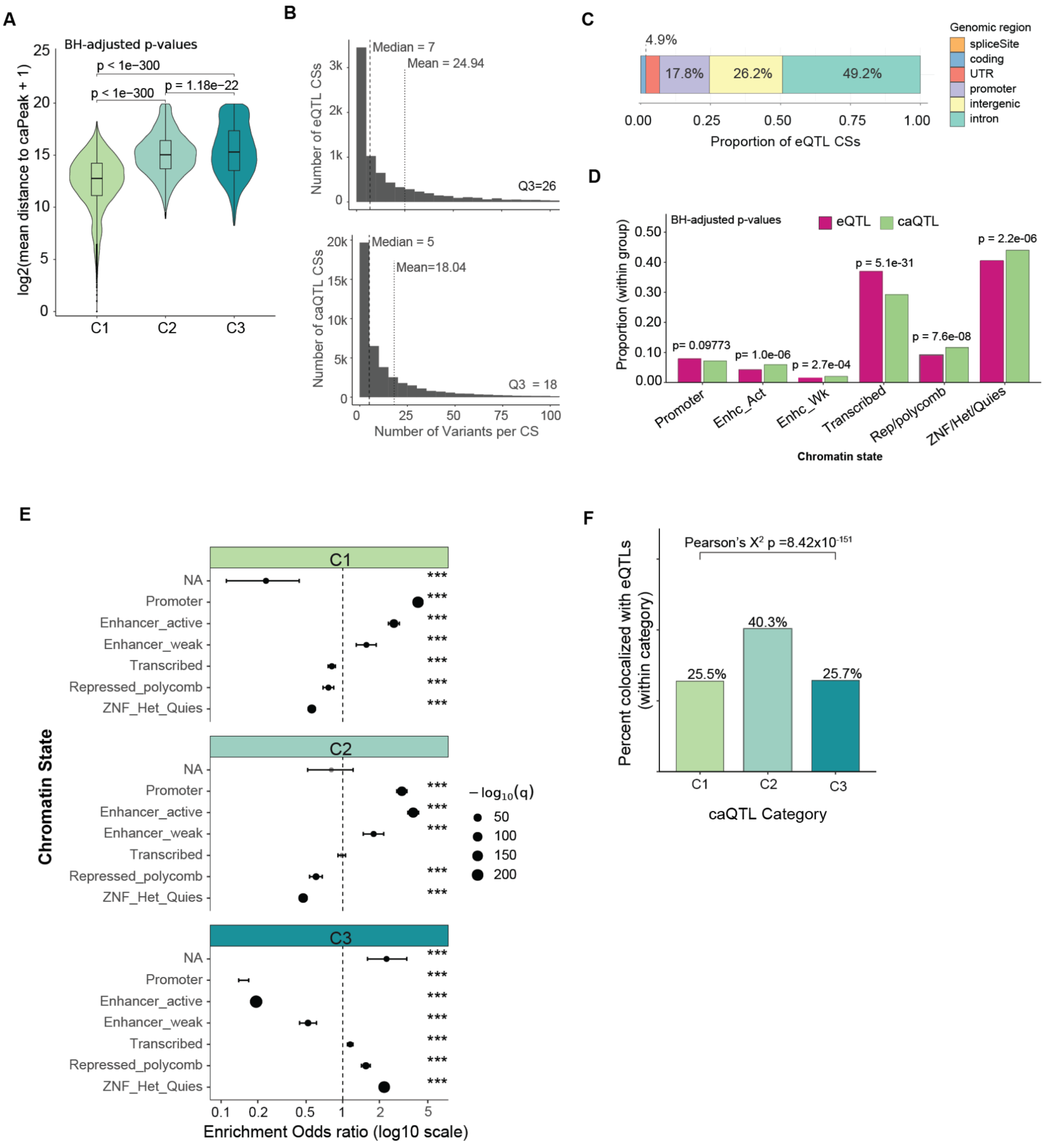

**Figure S4. Supporting analyses for differential regulatory architecture across molQTL modalities.** (**A**) Wilcoxon rank-sum test comparing the mean distance between caQTL credible sets (CSs) and their linked caPeaks; p-values were Benjamini-Hochberg (BH) adjusted. (**B**) Distribution of the number of variants per CS for each molQTL class. (**C**) Genomic annotation of eQTL CS variants. (**D**) Comparison of CS representation across ENCODE chromatin-state annotations between eQTLs and caQTLs (Pearson's chi-square test; BH-adjusted p-values). (**E**) Enrichment of CSs across ENCODE chromatin-state annotations within caQTL overlap categories (Fisher's exact test). Points show odds ratios (OR); horizontal lines indicate 95% confidence intervals. P-values were BH-corrected across chromatin states within each caQTL category; significance is denoted by asterisks. (**F**) Differences in eQTL colocalization rates across caQTL categories (chi-square test).

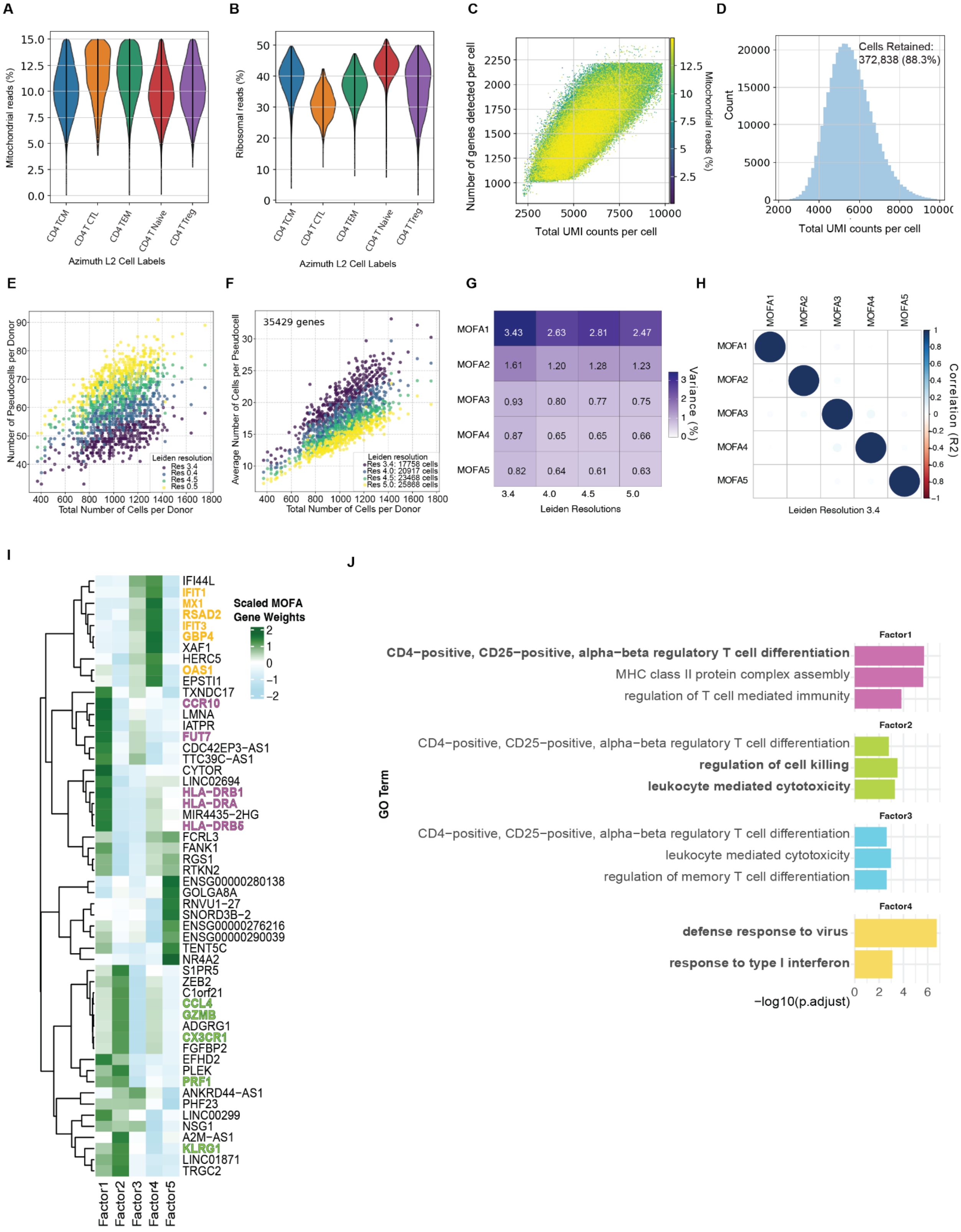
A
Mitochondrial reads (%)
Azimuth L2 Cell Labels
CD4 TCM
CD4 CTL
CD4 TEM
CD4 T Naive
CD4 T Treg
B
Ribosomal reads (%)
Azimuth L2 Cell Labels
C
Number of genes detected per cell
Total UMI counts per cell
Mitochondrial reads (%)
D
Cells Retained:
372,838 (88.3%)
Count
Total UMI counts per cell
E
Number of Pseudocells per Donor
Total Number of Cells per Donor
Leiden resolution
Res 3.4
Res 0.4
Res 4.5
Res 0.5
F
35429 genes
Average Number of Cells per Pseudocell
Total Number of Cells per Donor
Leiden resolution
Res 3.4: 17758 cells
Res 4.0: 20917 cells
Res 4.5: 23468 cells
Res 5.0: 25868 cells
G
MOFA1 3.43 2.63 2.81 2.47
MOFA2 1.61 1.20 1.28 1.23
MOFA3 0.93 0.80 0.77 0.75
MOFA4 0.87 0.65 0.65 0.66
MOFA5 0.82 0.64 0.61 0.63
3.4 4.0 4.5 5.0
Leiden Resolutions
Variance (%)
H
MOFA1 MOFA2 MOFA3 MOFA4 MOFA5
Correlation (R2)
Leiden Resolution 3.4
I
Scaled MOFA Gene Weights
IFI44L
IFIT1
MX1
RSAD2
IFIT3
GBP4
XAF1
HERC5
OAS1
EPSTI1
TXNDC17
CCR10
LMNA
IATPR
FUT7
CDC42EP3-AS1
TTC39C-AS1
CYTOR
LINC02694
HLA-DRB1
HLA-DRA
MIR4435-2HG
HLA-DRB5
FCRL3
FANK1
RGS1
RTKN2
ENSG00000280138
GOLGA8A
RNVU1-27
SNORD3B-2
ENSG00000276216
ENSG00000290039
TENT5C
NR4A2
S1PR5
ZEB2
C1orf21
CCL4
GZMB
ADGRG1
CX3CR1
FGFBP2
EFHD2
PLEK
PRF1
ANKRD44-AS1
PHF23
LINC00299
NSG1
A2M-AS1
KLRG1
LINC01871
TRGC2
Factor1 Factor2 Factor3 Factor4 Factor5
J
GO Term
Factor1
CD4-positive, CD25-positive, alpha-beta regulatory T cell differentiation
MHC class II protein complex assembly
regulation of T cell mediated immunity
Factor2
CD4-positive, CD25-positive, alpha-beta regulatory T cell differentiation
regulation of cell killing
leukocyte mediated cytotoxicity
Factor3
CD4-positive, CD25-positive, alpha-beta regulatory T cell differentiation
leukocyte mediated cytotoxicity
regulation of memory T cell differentiation
Factor4
defense response to virus
response to type I interferon
-log10(p.adjust)

**Figure S5. Supporting analyses for dynamic eQTL discovery and interpretation.** (**A–B**) Violin plots showing the distributions of (**A**) mitochondrial-read percentage and (**B**) ribosomal-read percentageacross CD4$^+$ T cell subtypes. (**C**) Scatter plot of genes detected per cell versus UMI counts per cell, colored by mitochondrial-read percentage. (**D**) Distribution of per-cell UMI counts after quality-control filtering. (**E**) Scatter plot comparing the number of cells per donor to the number of pseudocells per donor, colored by Leiden clustering resolution. (**F**) Scatter plot comparing the number of cells per donor to the mean number of cells per pseudocell, colored by Leiden clustering resolution. (**G**) Variance explained by MOFA models fit across Leiden resolutions. (**H**) Pairwise correlations between MOFA factors for the Leiden resolution 3.4 model. (**I**) Heatmap of top gene loadings for each MOFA factor. (**J**) Gene Ontology enrichment analysis of genes with the largest absolute MOFA loadings per factor (BH-adjusted p-values).

**A**

```
Query SNPs:
rs4786370 (chr16:3113898)
rs3760080 (chr16:3095818)

CEU Haplotypes:
               rs3760080
                  A      G
               ----------------
               C | 92    | 2     | 94    (0.475)
rs4786370      ----------------
               T | 54    | 50    | 104   (0.525)
               ----------------
                 146     52      198
               (0.737) (0.263)

               C_A: 92 (0.465)
               T_A: 54  (0.273)
               T_G: 50  (0.253)
               C_G: 2    (0.01)

          D': 0.919
          R2: 0.2719
          Chi-sq: 53.8304
          p-value: <0.0001

rs4786370(C) allele is correlated with rs3760080(A) allele
rs4786370(T) allele is correlated with rs3760080(G) allele
```

B

```
Query SNPs:
rs2272092 (chr7:150264645)
rs9691770 (chr7:150254163)

CEU Haplotypes:
               rs9691770
                  A       G
               -----------------
               C | 69    | 0         | 69        (0.348)
rs2272092      -----------------
               T | 0      | 129      | 129       (0.652)
               -----------------
                   69      129       198
                (0.348) (0.652)

               T_G: 129 (0.652)
               C_A: 69 (0.348)
               C_G: 0 (0.0)
               T_A: 0 (0.0)

               D': 1.0
               R2: 1.0
               Chi-sq: 198.0
               p-value: <0.0001

rs2272092(C) allele is correlated with rs9691770(A) allele
rs2272092(T) allele is correlated with rs9691770(G) allele
```

**Figure S6. LD-based variant linkage queries. (A)** Linkage disequilibrium between rs3760080 and rs4786370. **(B)** Linkage disequilibrium between rs9691770 and rs2272092. LD was evaluated in the 1000 Genomes CEU reference population (Utah Residents with Northern and Western European ancestry).

**A**

| Fold | Train | Validation | Test |
|---|---|---|---|
| 0 | chr2, chr4, chr5, chr7, chr9, chr10, chr11, chr12, chr13, chr15, chr16, chr17, chr18, chr19, chr21, chr22 | chr8, chr20 | chr1, chr3, chr6, chr14 |
| 1 | chr1, chr3, chr4, chr5, chr6, chr7, chr10, chr11, chr13, chr14, chr15, chr18, chr19, chr20, chr21, chr22 | chr12, chr17 | chr2, chr8, chr9, chr16 |
| 2 | chr1, chr2, chr3, chr5, chr6, chr8, chr9, chr10, chr13, chr14, chr16, chr17, chr18, chr19, chr20, chr21 | chr22, chr7 | chr4, chr11, chr12, chr15 |
| 3 | chr1, chr2, chr3, chr4, chr7, chr8, chr9, chr11, chr12, chr13, chr14, chr15, chr16, chr17, chr19 | chr6, chr21 | chr5, chr10, chr18, chr20, chr22 |
| 4 | chr1, chr2, chr3, chr4, chr5, chr6, chr8, chr9, chr11, chr12, chr14, chr15, chr16, chr20, chr22 | chr10, chr18 | chr7, chr13, chr17, chr19, chr21 |

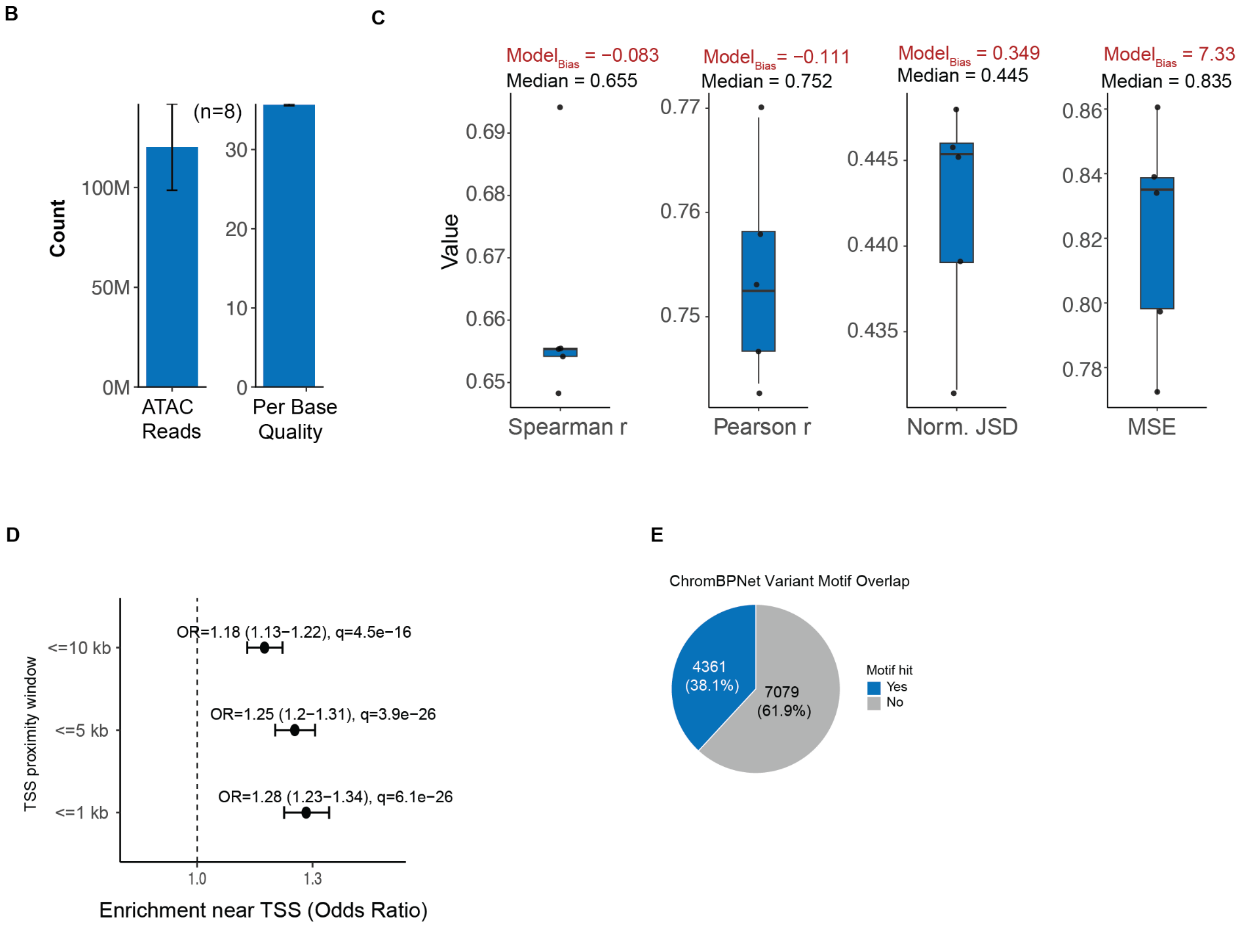

**Figure S7. ChromBPNet model training, QC, and motif-overlap summary.** (**A**) Chromosome-level partitioning across training, validation, and test sets for each of five cross-validation folds. (**B**) Sequencing-depth and base-quality summary for the eight ATAC-seq samples used for ChromBPNet training: (1) total aligned ATAC-seq reads per sample and (2) mean base quality score per sample. Bars show the mean across samples (n = 8); error bars indicate ± SD across samples. (**C**) Bias-factorized ChromBPNet performance across cross-validation folds. Boxplots show per-fold metrics including Spearman's ρ, Pearson's r, normalized Jensen–Shannon divergence (JSD), and mean squared error (MSE); red points indicate the corresponding metrics for the bias-only model. (**D**) Proportion of significantly scored ChromBPNet variants that overlap at least one motif instance, relative to the total number of significantly scored variants. (**E**) Enrichment of significantly scored ChromBPNet variants in windows proximal to the TSS of any gene (≤1 kb, ≤5 kb, ≤10 kb). Points show odds ratios (OR); horizontal lines indicate 95% confidence intervals. P-values were BH-corrected across windows.

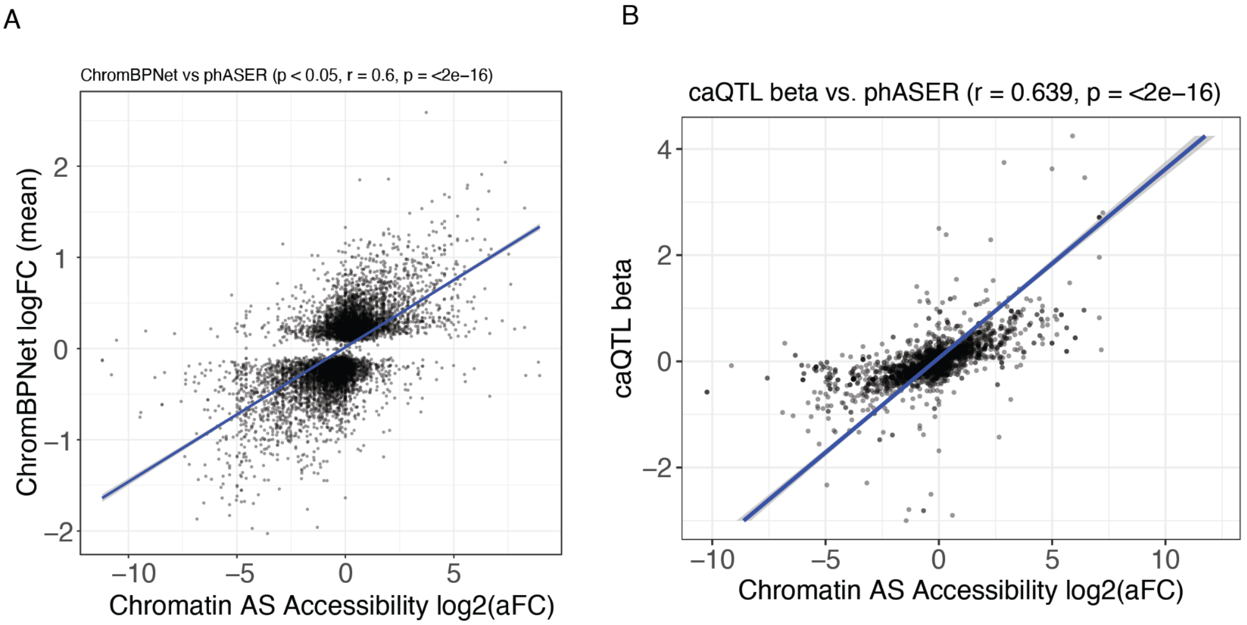

**Figure S8. Concordance between ChromBPNet predictions, allelic chromatin accessibility, and caQTL effect sizes**. (**A**) Comparison of ChromBPNet-predicted allelic chromatin accessibility effects (predicted log fold change, logFC) with empirically observed allele-specific chromatin accessibility log fold change (logFC) at the corresponding loci. (**B**) Comparison of caQTL effect sizes (β) with allele-specific chromatin accessibility log fold change (logFC) at the corresponding loci.

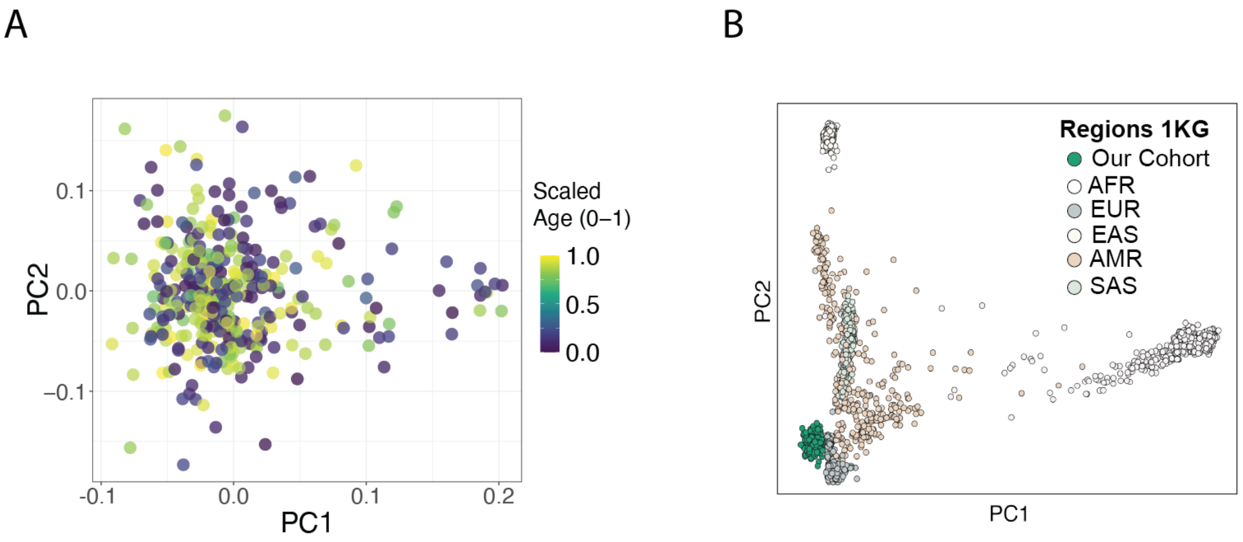

**Figure S9. Genetic ancestry structure of the study cohort.** (**A**) Principal component analysis (PCA) of genotyped samples after quality control, colored by scaled ages. (**B**) PCA of the study cohort projected alongside the 1000 Genomes reference panel to assess genetic ancestry structure.

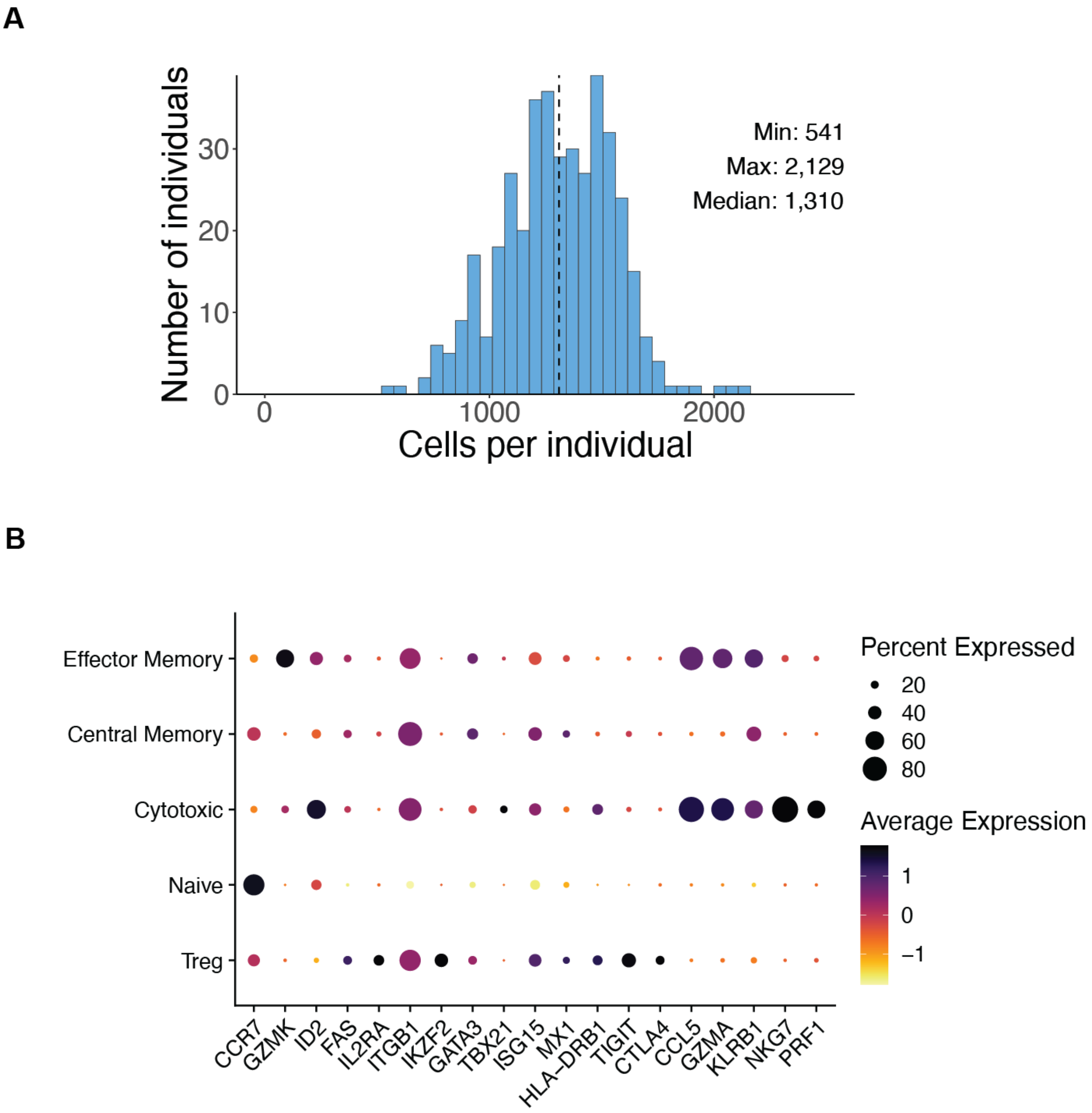

**Figure S10. scRNA-seq quality control and validation of CD4$^+$ T cell subtype labels.** (**A**) Distribution of cells per individual after scRNA-seq quality control and filtering. (**B**) Dot plot of canonical marker expression across Azimuth-labeled CD4$^+$ T cell subtypes.

A

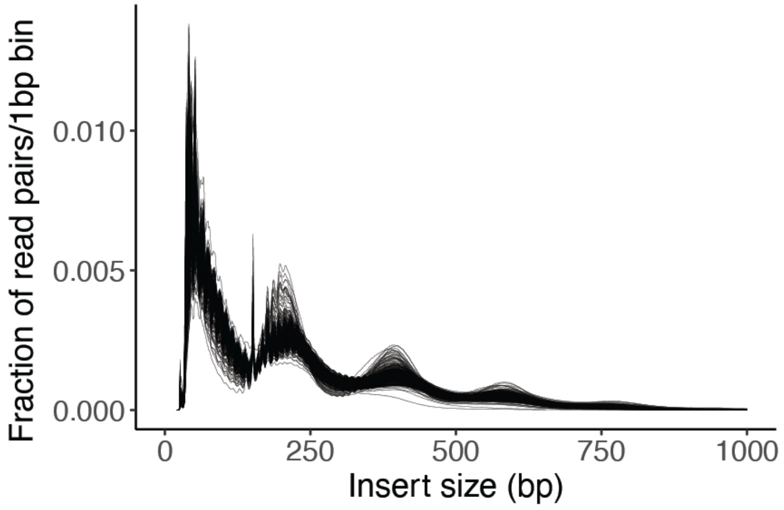

B

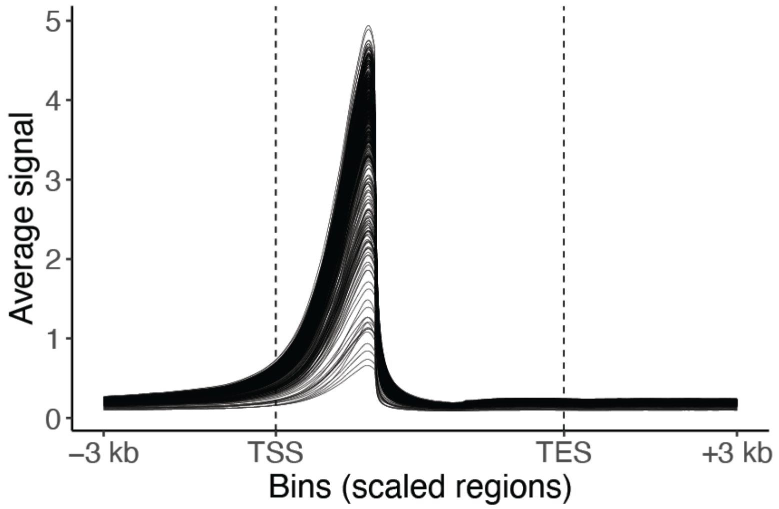

**Figure S11. Figure S4. ATAC-seq fragment-length distribution and TSS enrichment.**

(**A**) Distribution of ATAC-seq fragment (insert) sizes across all samples (showing expected nucleosomal periodicity). (**B**) Aggregated enrichment of ATAC-seq signal around transcription start sites (TSS), assessed using deepTools profile/heatmap summaries.

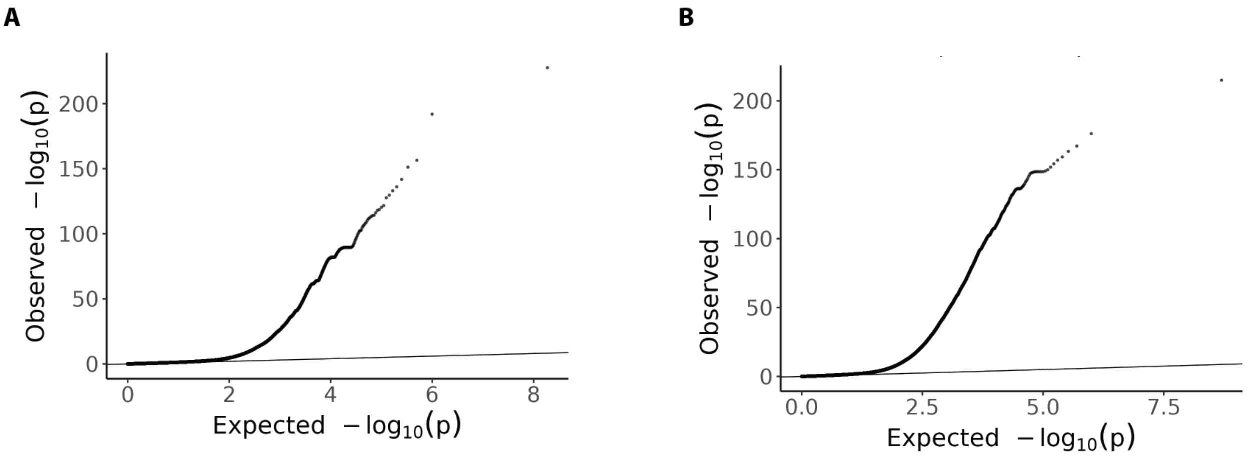

**Figure S12. molQTL discovery QQ plots. (A)** Quantile–quantile (QQ) plot of nominal p-values from cis-eQTL mapping. **(B)** Quantile–quantile (QQ) plot of nominal p-values from cis-caQTL mapping.
